# Rapid submillimeter QSM and R_2_^*^ mapping using interleaved multi-shot 3D-EPI at 7 and 3 Tesla

**DOI:** 10.1101/2023.12.29.23300637

**Authors:** Rüdiger Stirnberg, Andreas Deistung, Jürgen R. Reichenbach, Monique M. B. Breteler, Tony Stöcker

## Abstract

**Purpose:** To explore the high signal-to-noise ratio (SNR) efficiency of interleaved multi-shot 3D-EPI for fast and robust high-resolution whole-brain quantitative susceptibility (QSM) and 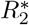 mapping at 7T and 3T.

**Methods:** Single- and multi-TE segmented 3D-EPI is combined with conventional CAIPIRINHA undersampling for up to 72-fold effective gradient echo (GRE) imaging acceleration. Across multiple averages, scan parameters are varied (e.g. dual-polarity frequency-encoding) to additionally correct for *B*_0_-induced artifacts, geometric distortions and motion retrospectively. A comparison to established GRE protocols is made. Resolutions range from 1.4mm isotropic (1 multi-TE average in 36s) up to 0.4mm isotropic (2 single-TE averages in approximately 6 minutes) with whole-head coverage.

**Results:** Only 1-4 averages are needed for sufficient SNR with 3D-EPI, depending on resolution and field strength. Fast scanning and small voxels together with retrospective corrections result in substantially reduced image artifacts, which improves susceptibility and 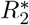 mapping. Additionally, much finer details are obtained in susceptibility-weighted image projections through significantly reduced partial voluming.

**Conclusion:** Using interleaved multi-shot 3D-EPI, single-TE and multi-TE data can readily be acquired 10 times faster than with conventional, accelerated GRE imaging. Even 0.4mm isotropic whole-head QSM within 6 minutes becomes feasible at 7T. At 3T, motion-robust and distortion-free 0.8mm isotropic whole-brain QSM and 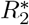 mapping in less than 7 minutes becomes clinically feasible. Stronger gradient systems may allow for even higher effective acceleration rates through larger EPI factors while maintaining optimal contrast.

## 1 Introduction

Quantitative susceptibility mapping (QSM) and mapping of the effective transverse relaxation rate 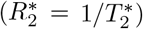 provide valuable quantitative information about biological tissue in vivo. For instance, voxel-wise information on magnetic susceptibility, *χ*, and 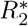 allows drawing spatially resolved conclusions on deep gray matter iron content^1, 2^ and white matter myelination.^3^ Using *χ* and 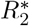 together even allows separating the two sources on a sub-voxel level.^4–6^

Magnitude and phase MR images for 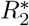 mapping and QSM are usually acquired using volumetric (3D) spoiled gradient echo (GRE) imaging. Often, relatively short repetition times (*TR ≪ T*_2_) are combined with low flip angle excitations to maximize signal for the longitudinal relaxation time (*T*_1_) of a specific tissue. Within each TR, a different phase-encoded k-space line is acquired using a frequency-encoding gradient pulse.

For optimal 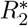 and susceptibility contrast, the echo time (TE) should be on the order of the average tissue 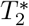. For 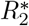 mapping, a reasonable range of short-to-long TEs should be sampled. Both result in rather long TR and, depending on the desired voxel size, can lead very long acquisition times (TA) and related artifacts (motion, field fluctuations, etc.), even with state-of-the-art imaging acceleration. Increasing image acceleration beyond that through advanced parallel imaging (e.g. [^7^]) can become cumbersome, as it typically requires offline image reconstruction with the exact knowledge of actual gradient trajectories. Otherwise, typical compromises to shorten TA include anisotropic voxels, reduced slab coverage in slice direction or the use of shorter, but suboptimal TE and TR.

Instead of sacrificing contrast-to-noise ratio (CNR), it is desirable to keep TE and TR long, and reduce TA through faster scanning with highest possible efficiency (signal-to-noise ratio, SNR, per unit scan time). Previous work has used echo planar imaging (EPI) to acquire GRE data more efficiently for rapid QSM (e.g. [^8, 9^]). Such approaches, however, did not consider multi-shot segmentation and relied only on single-TE acquisitions, preventing high-resolutions as well as multi-TE QSM or 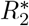 mapping. On the other hand, interleaved multi-shot 3D-EPI has already been utilized successfully for high-resolution 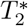- and susceptibility-weighted imaging as well as multiparametric mapping, including 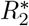 mapping, at 3T and 7T.^10–12^ Such approaches combined with QSM have vast neuroscientific potential (e.g. [^13^]).

We show that combining *R*-fold CAIPIRINHA^14^ undersampling with an EPI factor *N >* 1 allows for significant imaging acceleration (up to *N × R*-fold) making retrospective image-based corrections feasible, paving the way to imaging at ultra-high isotropic resolutions. We demonstrate particularly efficient whole-head QSM and 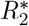 mapping using single-TE and multi-TE interleaved multi-shot 3D-EPI at 7T and 3T with isotropic resolutions as high as 400 microns.

## 2 Theory

### 2.1 SNR efficiency of gradient echo imaging

In appendix A it is shown that steady-state GRE imaging (of a monoexponential free induction decay) is most SNR-efficient, when the Ernst angle is used and the signal is sampled throughout a particularly long TR (eq. (5)):

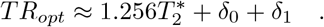

Here, *δ*_0_ and *δ*_1_ denote void times before and after signal sampling, respectively (see fig. 9 A,B,D). A long TR is beneficial for acquisitions with rather long TE as required for optimal 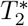- and susceptibility-weighting. If signal acquisition is split throughout the TR (e.g. multiecho GRE, see fig. 9 A,C,E), the SNR-optimal TR becomes even longer, according to appendix B. In both cases, however, SNR efficiency drops relatively quickly, when shorter TRs are used (see fig. 1).

**Figure 1:**
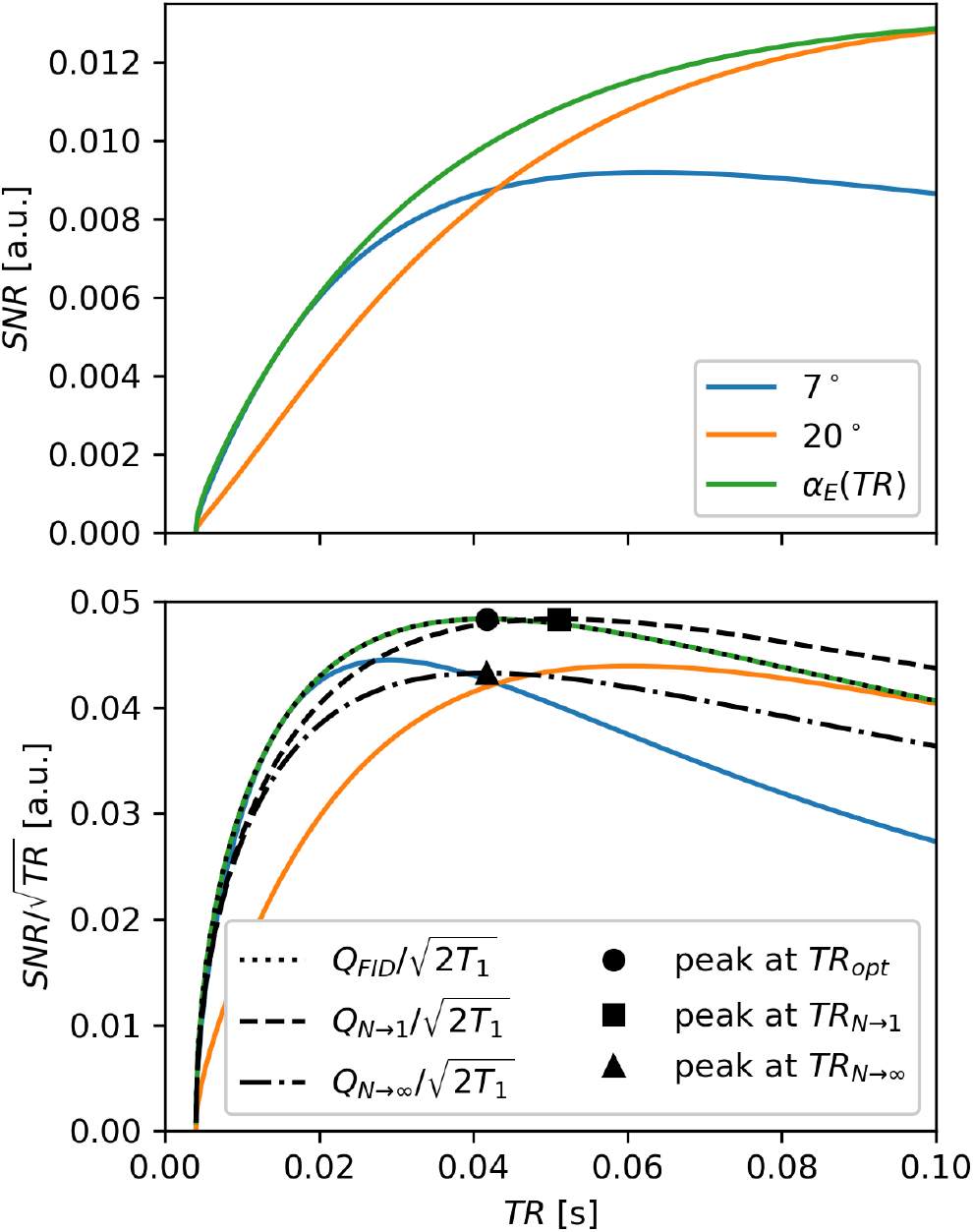
SNR (top) and SNR efficiency (bottom) of spoiled GRE as a function of TR for different flip angles (solid lines) and the Ernst angle (*α*_*E*_(*TR*)) assuming *T*_1_ = 2.0s and 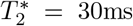. Bottom: approximated efficiency curves and peaks for a single FID (dotted) and the extremal cases of the multi(*N*)-echo model (dashed: *N →* 1, dash-dotted: *N → ∞*). All curves are only defined for *TR >* 4ms (the sequence void time assumed here, e.g. for phase-encoding). The multi-echo model assumes that a fraction of the overall sampling window (here: 20%) is not sampled.

While there are different ways to implement GRE with long TR in practice, a single phase-encoding step per TR typically results in long TA. Here, three cases are considered, the first two of which are commonly used in QSM:

#### Single-echo GRE

A single-TE GRE would have to use a very small frequency-encode bandwidth to make SNR-efficient use of the largest possible sampling window. This would lead to overly strong susceptibility-induced artifacts, rather long TA and, in most feasible cases, very high SNR. Typically, a less SNR-efficient but feasible compromise is chosen (see single-TE GRE experiment 1a).

#### Multi-echo GRE

Using *N* GRE readouts with approximately *N* -fold increased bandwidth (to fit them into the same TR) would reduce susceptibility-induced artifacts while providing multi-echo information. The *N* individual images would have 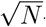 -fold increase in noise. However, if used jointly, e.g. in a derived susceptibility map, all images together could provide comparable SNR to the single-TE GRE. Nevertheless, TA remains long, or spatial resolution must be compromised for shorter TA (see multi-TE GRE experiment 1b).

#### Multi-shot 3D-EPI

In contrast to the two previous approaches, TA can be strongly reduced by acquiring multiple k-space lines per TR. Then, TA reduces by the EPI factor *N* (a.k.a. echo train length). While SNR is also reduced by 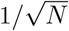 compared to the single-TE GRE case, SNR efficiency is still as high. This means that approximately the same SNR can be regained by acquiring and averaging *N* (complex-valued) images in the same TA. However, depending on the application, the SNR of *M < N* averages may already suffice and TA would be only a fraction *M/N* compared to the two GRE cases. For this strategy, it typically makes sense to chose *N* as large as possible, i.e. maximize frequency-encode bandwidth (minimize echo spacing, *ESP*) and chose the largest EPI factor that fits into TR while meeting TE at the k-space center line (see high-resolution single-TE EPI experiments 1a and 2a).

### 2.2 Interleaved multi-shot 3D-EPI

Taking advantage of its high SNR efficiency, an interleaved multi-shot 3D-EPI sequence with CAIPIRINHA sampling^14, 15^ is considered in this work for maximum acceleration capabilities along the primary and secondary phase-encode directions (*y* and *z* w.l.o.g.). Using moderately high parallel imaging undersampling, *R*_*y*_ *· R*_*z*_ ≤ 6, and a small number of averages, *M*, still quite high effective acceleration rates can be achieved using a large EPI factor, *N* :

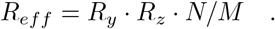

Susceptibility-induced artifacts along the frequency-encode direction (*x*, w.l.o.g.) are negligible as very high bandwidths are typically used. The interleaved EPI readout results in a steady, but lower bandwidth along *y*. Still, with high segmentation (*S*), actual phase-encode bandwidths,

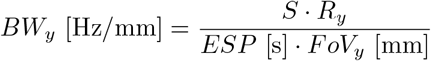

become comparable to the frequency-encode band-widths of typical GRE protocols (cf. table 1). Here, *FoV*_*y*_ refers to the phase-encode field-of-view.

**Table 1:**
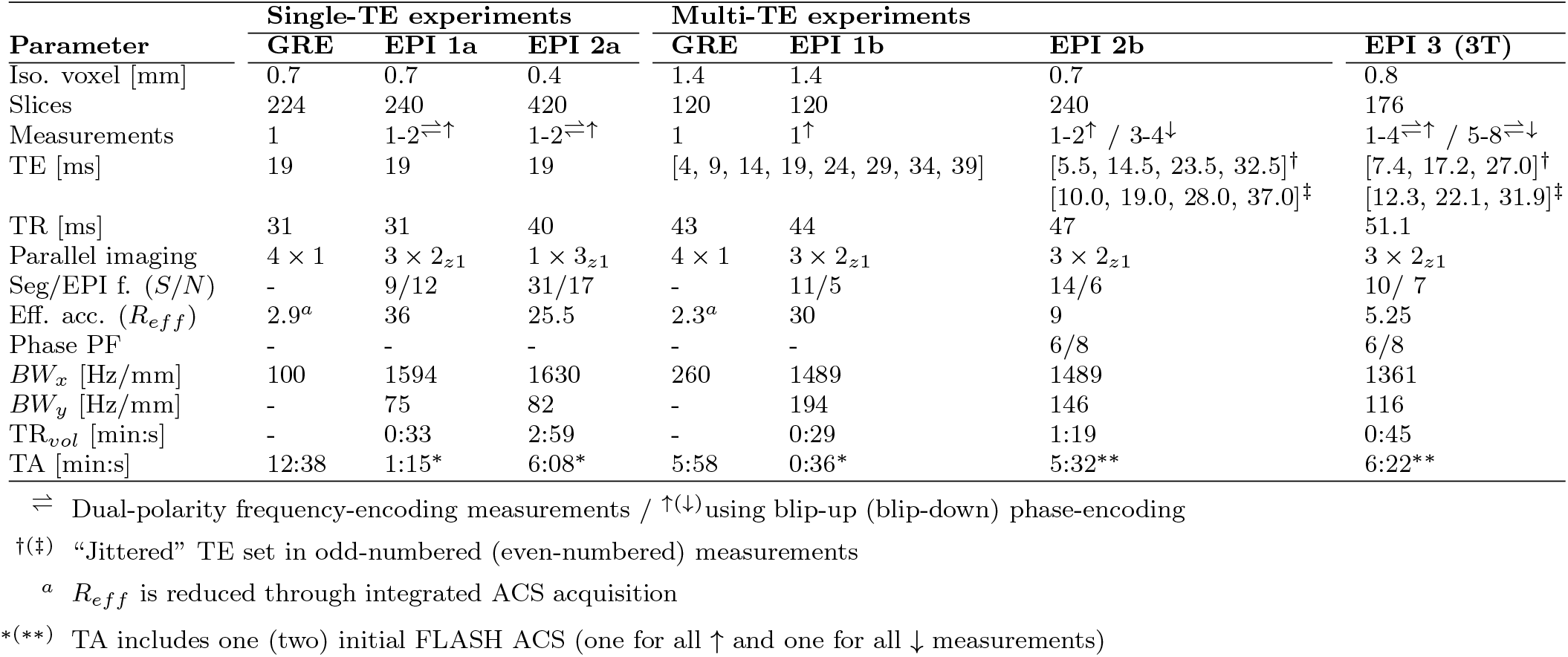
Protocol parameters.

Finally, by adding phase-encode rephasers regularly between every few EPI echoes, a multi-TE EPI readout is implemented^12^ that trades multi-echo information for TA, but still performs at comparable SNR efficiency (see multi-TE experiment 1b). If sufficiently short multi-TE EPI readouts cannot be realized with a practical EPI factor, we utilize “jittered” TEs across measurements (see high-resolution multi-TE experiments 2b and 3).

### 2.3 Retrospective corrections and dual-polarity frequency-encoding

Unlike a single, long GRE acquisition, *M* fast EPI acquisitions can be utilized, for instance, to retrospectively correct for inter-volume motion before averaging. Likewise, local phase changes over time (e.g. due to frequency drift or motion) can be matched before averaging. If the phase encode polarity is changed for at least one measurement, it is also possible to estimate and correct geometric distortions along the phase-encode direction before averaging.

Another option is to invert the EPI frequency-encode polarity in every other scan (dual-polarity frequency-encoding). When reordered, two complete sets of cohesive, single-polarity k-space data according to fig. 2C,D could be obtained from two EPI measurements according to fig. 2A,B: one acquired with only positive (red lines) and one with only negative frequency-encode polarity (blue lines). The two hypothetical fly-back images would, for instance, not suffer from Nyquist ghosts, even if acquired with a systematic *k*_*x*_ shift between positive and negative lines. If navigator-based phase correction were already performed, as in this work, the residual phase discrepancy would become negligible: The k-space data could be averaged to yield a high-SNR image without Nyquist ghosts.

**Figure 2:**
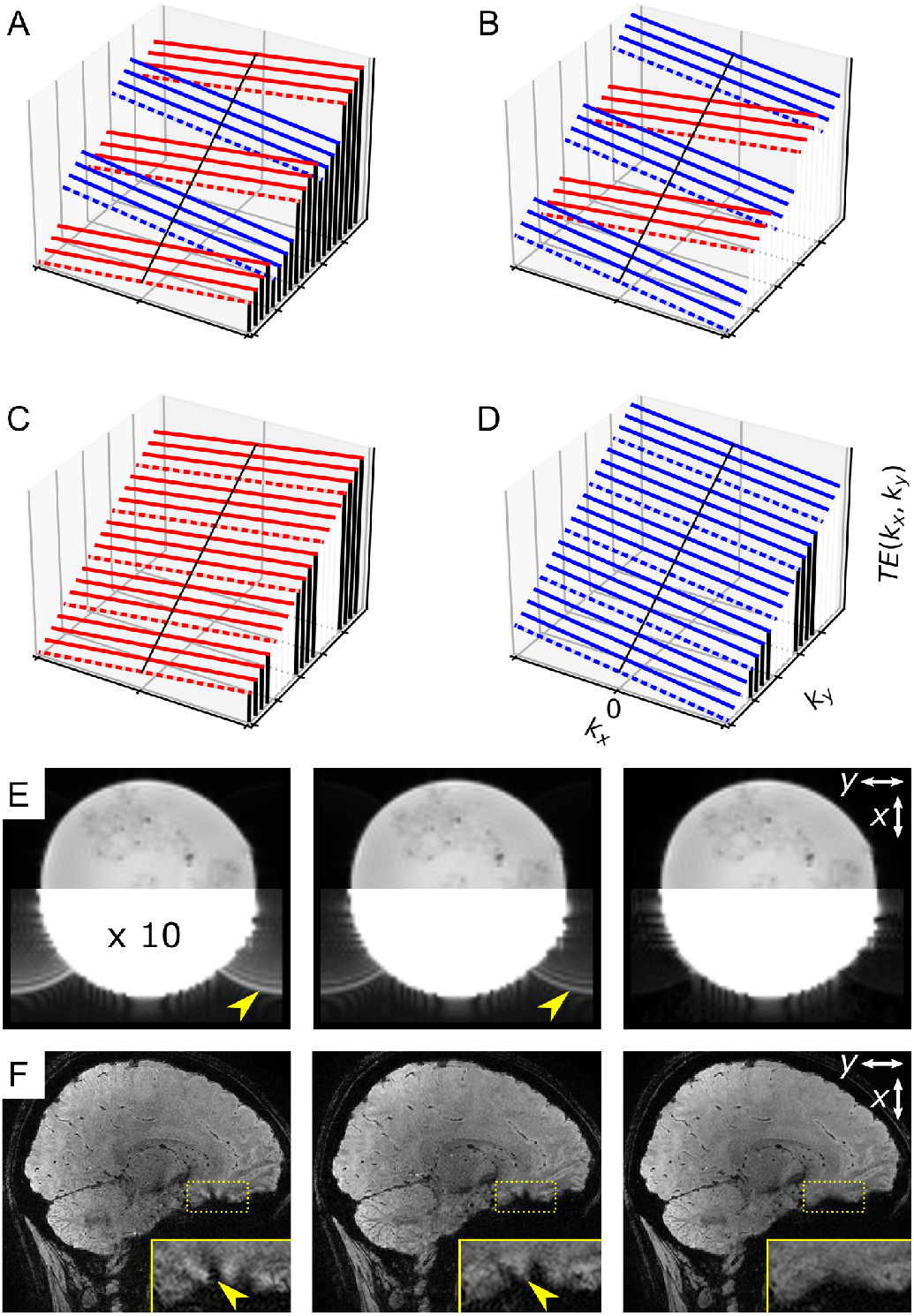
TE (phase error) vs. *k*_*x*_ × *k*_*y*_ for dual-polarity acquisition (A,B) and hypothetical reordering (C,D) of interleaved multi-shot EPI with ETS (*S* = 4, *N* = 5, dashed: first interleave, red/blue: pos./neg. frequency-encoding polarity). Mitigated Nyquist ghosts (E) and segmentation artifacts (F) after dual-polarity averaging of images according to (A,B). Spherical phantom intensities scaled by a factor of 10 show low-intensity Nyquist ghosts before averaging (E, 3T, no segmentation). Magnified insets in (F) show complementary segmentation artifacts before averaging and a reduced, homogeneous dropout area after averaging (7T, *N* = 12, *S* = 9, 3 *×* 2_*z*1_). *x*: frequency-encode, *y*: phase-encode.

#### 2.3.1 Mitigation of segmentation artifacts

We propose dual-polarity frequency-encoding to mitigate off-resonance-induced segmentation artifacts in interleaved multi-shot EPI, as demonstrated in fig. 2F. While echo time shifting (ETS)^16^ is already implemented for a smooth, linear TE evolution along *k*_*y*_ at *k*_*x*_ = 0, off-resonance-induced phase jumps still remain between echo sections (contiguous red and blue sections in fig. 2A,B) for all *k*_*x ≠*_ 0. The two hypothetical fly-back images according to fig. 2C,D would be free of segmentation artifacts. An off-resonant voxel would only be displaced in opposite directions along *x* (susceptibility-induced geometric distortions). With large frequency-encode bandwidths, as used in EPI, the displacements are usually well below the nominal voxel size for typical off-resonance frequencies in the brain. Consequently, averaging the two reordered k-space sets would yield an image free of segmentation artifacts. Blurring along *x* would occur only where voxels are extremely off-resonant (cf. Supporting information figure S1).

#### 2.3.2 Dual-polarity averaging

As the Fourier transform between k-space and image space is linear, Nyquist ghosts and segmentation artifacts can be mitigated as described without the need to reorder positive and negative frequency-encode lines, and averaging can be performed in the image domain instead of the k-space domain: From two complex-valued dual-polarity images with residual Nyquist ghosts and segmentation artifacts, one retrospectively averaged image with eliminated Nyquist ghosts and mitigated segmentation artifacts is obtained, as demonstrated in fig. 2E and F.

## 3 Methods

### 3.1 Experiments

A custom skipped-CAIPI 3D-EPI (SC-EPI) sequence^15^ that optionally inverts the frequency-encode polarity in every other measurement, was implemented at 7T (Siemens Healthineers, MAGNETOM 7TPlus) and at 3T (MAGNETOM Prisma). Both scanners are equipped with a high-performance gradient system (70 mT/m, 200 T/m/s at 7T; 80 mT/m, 200 T/m/s at 3T). At 7T, a 32-channel head receive coil surrounded by a single-channel (circular-polarized) transmit coil was used. At 3T, the 52 head elements of a 64 channel head-neck receive coil were used. Image reconstruction was performed using the vendor-based implementation (“IcePAT”) that applies 3D GRAPPA kernels and a prescan-based, virtual reference coil combination.^17^

As a reference at 7T, GRE protocols were set up as listed below based on the recent multi-centre, multi-vendor UK7T reproducibility study.^18^ All 7T scans were performed in sagittal slice orientation (*x*: head-feet, *y*: posterior-anterior) using non-selective narrow bandwidth RF pulses to suppress fat signal.^19^ The vendor-provided 3D-GRE sequence was adapted to use the same image reconstruction as the SC-EPI. However, in accordance with the UK7T protocols for the Siemens MAGNETOM Terra 7T scanner,^20^ all GRE scans used a zero CAIPIRINHA shift and “integrated” GRAPPA autocalibration scans (ACS). Flow compensation (along *x* and *z*) was only active for the single-TE GRE. The multi-TE GRE used a monopolar readout.

All EPI scans used fast initial FLASH ACS^21^ and no flow compensation. The rapid single-TE EPI used integrated phase navigators for shot-by-shot phase correction, whereas all other EPI protocols used a one-time initial phase navigator for more efficient but static phase correction.^15^

The *x* × *y* field-of-view (FoV) was 224mm × 224mm for all scans, except for the multi-TE GRE (269mm × 219mm) and the high-resolution single-TE EPI (205mm × 205mm). The nominal flip angle was 15° for all scans, except for the high-resolution multi-TE EPI (18°). Only the latter used a phase partial Fourier (PF) factor of 6/8 and both blip-up (↑) and blip-down (↓) phase-encoding. Further protocol details are summarized in table 1.

All scans were acquired with written informed consent from three subjects in accordance with the local ethics regulations. The following use cases of whole-head SC-EPI are examined:

1. Ultra-fast QSM and 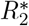 mapping at 7T:
  a. Single-TE at 0.7mm isotropic resolution
  b. Multi-TE at 1.4mm isotropic resolution
2. High-resolution QSM and 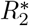 mapping at 7T:
  a. Single-TE at 0.4mm isotropic resolution
  b. Multi-TE at 0.7mm isotropic resolution
3. High-resolution QSM and 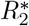 mapping at 3T: Multi-TE at 0.8mm isotropic resolution

Compared to the UK7T-based GRE protocols, experiments 1a and 1b provide ten times shorter acquisition times. Experiments 2a and 2b provide 1/5 and 1/8 of the respective voxel volumes, while still being twice as fast.

Experiment 3 transfers the 7T high-resolution multi-TE EPI protocol to 3T for routine application in the Rhineland Study – a long-term, large-scale study in the general population^22^ (https://www.rheinland-studie.de/en/). Unlike the 7T SC-EPIs, single-oblique axial slices (*x*: left-right, *y*: anterior-posterior) were acquired using slab-selective, binomial-121 water excitation. A 216mm × 216mm *x* × *y* FoV is used. A 150mm thick saturation slab, positioned below and parallel to the imaging slab and applied before each excitation, suppressed in-flowing blood signal. At 3T, both dual-polarity frequency-encoding and ↑ & ↓ phase-encoding were combined with two sets of “jittered” TEs, yielding *M* = 8 measurements in total, each with a *TR*_*vol*_=45s. Further parameters are listed in table 1. At 3T, the volunteer was asked to lie still during a first pass, and instructed to perform one realistic and one exaggerated motion during a second pass of the entire scan (see Supporting information Fig. S2).

### 3.2 Preprocessing

If only a single SC-EPI image was acquired per TE, no preprocessing was performed (experiment 1b). Otherwise, an image-based preprocessing pipeline was implemented using standard software (e.g. FSL^23^) and custom basic python code (e.g. “phase matching” and “PF compensation”) as outlined in fig. 3.

**Figure 3:**
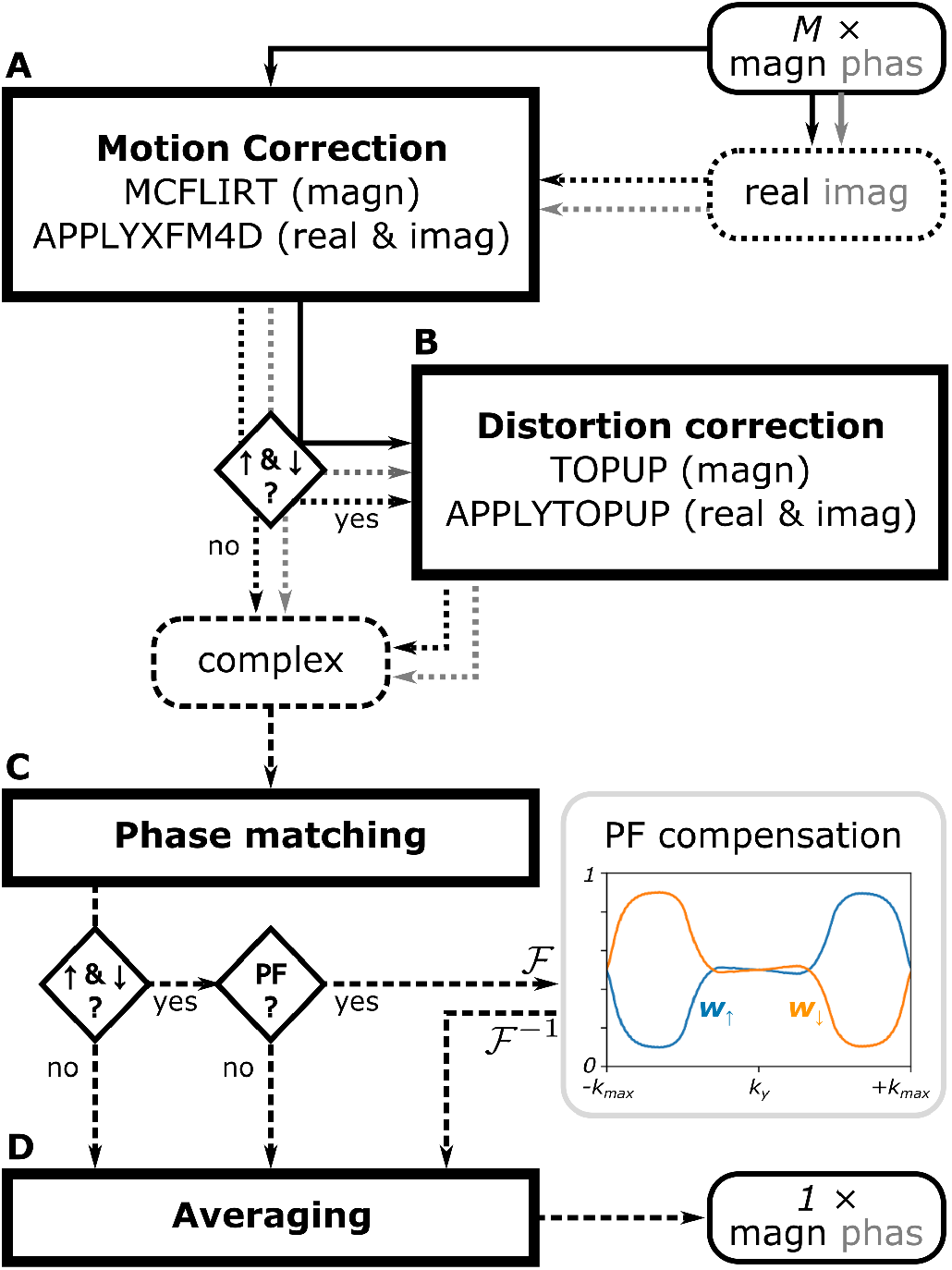
Preprocessing pipeline that turns *M* complex-valued native SC-EPI images into one complex-valued high-SNR image per TE. The input images are retrospectively corrected for motion (A), geometric distortions (B) and phase changes (C). Finally, complex-valued averaging (D) is carried out to mitigate residual Nyquist ghosts and segmentation artifacts (if dual-polarity frequency-encoding was applied) and/or to compensate for partial Fourier (PF) blurring (if ↑ & ↓ phase-encoding was applied). Different complex image *ℱ* and *ℱ*^*−*1^: data formats are represented by line type. and: Fast Fourier transforms to k-space and back to image space.

The minimal pipeline in this work consisted of motion correction (A), using the mcflirt and applyxfm4D methods implemented in FSL, and phase matching (C) before data averaging (experiments 1a and 2a). In an extended pipeline, the ↑ & ↓ phase-encoded images were used to estimate and correct for susceptibility-induced geometric distortions (B), using the topup and applytopup methods implemented in FSL (experiments 2b and 3). If dual-polarity frequency-encoding was applied (all experiments, except 1b and 2b), the final averaging step (D) also corrected for residual Nyquist ghosts and segmentation artifacts. If phase partial Fourier sampling (PF) was combined with ↑ & ↓ phase-encoding (experiments 2b and 3), final averaging also compensated for PF-related blurring through appropriate k-space weighting (see below).

#### 3.2.1 Phase matching

Local phase changes over *M* repeated measurements were accounted for by matching the low spatial frequency contributions to the phase with respect to the first measurement. For single-TE, this was carried out by subtracting a low-pass-filtered version of the phase changes (computed via the Hermitean-inner-product followed by a 2 × 2 × 2-voxel Gaussian kernel smoothing of the real and imaginary images). For multi-TE data, the actual frequency changes over measurements were estimated as the changes of the phase difference between the first two rephased TEs divided by the TE spacing. Numerical integration of the low-pass-filtered frequency changes across measurements, multiplied with the respective TEs yielded consistent phase changes to be subtracted. For both single-TE and multi-TE data, this phase matching strategy keeps the high spatial frequency noise of the original complex-valued images unaltered while the image phases are aligned.

#### 3.2.2 Partial Fourier compensation

When phase PF sampling is combined with *↑ & ↓* phase-encoding, the *↑* and *↓* data, each collected *M/*2 times, are zero-filled at opposite k-space ends. Therefore, actual data is acquired *M/*2 times at each end and *M* times in the overlapping k-space section. Here, the images ded not require individual PF reconstructions as compared to [^24^]. Instead, they were averaged using appropriate k-space weights in the style of [^24^]. For our data we found that smooth weights *w*_*↑*_(*k*_*y*_) and *w*_*↓*_(*k*_*y*_) resulted in better retrospective PF compensation than hard-coded weights (0, 0.5, 1) with sharp transitions. This is primarily due to the tapering involved in the preceding, zero-filled PF reconstruction. We derived the weights from the ↑ and ↓ k-space magnitudes themselves as

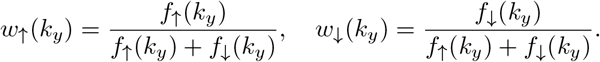

Herein, the high-SNR k-space magnitude functions *f*_*↑/↓*_ were obtained by root-sum-of-squares combination of the actual k-space signals *s*_*↑/↓*_ across all *k*_*x*_, *k*_*z*_, *TE*_*j*_ and repeated measurements (*m*),

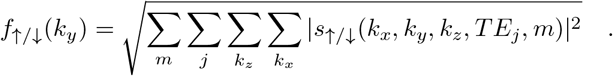

The resulting PF compensation weights were therefore consistent for all averages and TEs (examples shown in fig. 3).

#### 3.2.3 Complex-valued non-local means denoising

As a final, optional step, we applied a complex-valued customization of a non-local means (NLM) denoising algorithm.^25^ Here, the ANTsPy^26^ implementation was applied separately on the high-pass-filtered real and imaginary components achieved by subtracting a Gaussian-smoothed baseline (4 × 4 × 4-voxel kernel). Air voxels were excluded from denoising (first image magnitude quintile). Afterwards, the smooth baseline, assumed to have very little low-frequency noise, was added back to the denoised high-frequency components.

### 3.3 SWI, QSM and 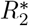 mapping

Susceptibility-weighted images^27^ were computed from the preprocessed magnitude and phase images considering all TEs using CLEAR-SWI^28^ (https://github.com/korbinian90/CLEARSWI.jl). For all SWI data, minimum intensity projections (MIP) across 5.6mm axial slabs are also presented in this work.

For QSM, the phase images for each measurement and TE were unwrapped using a 3D best path algorithm^29^ divided by 2*π · TE*_*j*_[s] to obtain the Larmor frequency variation in Hz. The latter were combined across TEs for multi-TE data (*TE*_*j*_ *≤* 19ms at 7T, all *TE*_*j*_ at 3T). Background field contributions were removed using Variable-radius Sophisticated Harmonic Artifact Reduction for Phase data (V-SHARP).^30, 31^ Susceptibility mapping was performed based on the V-SHARP-processed frequency maps using Homogeneity Enabled Incremental Dipole Inversion (HEIDI).^32^ All susceptibility values were referenced to the average susceptibility of brain tissue within the FoV.^33^

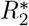 mapping was performed on all multi-TE magnitude images using the NUMerical Algorithm for Realtime 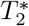 mapping (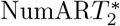)^34^ (https://github.com/korbinian90/MriResearchTools.jl).

## 4 Results

Figure 4 shows several views of the single-TE magnitude data at 7T and corresponding SWIs and MIPs. The preprocessing of both EPI scans consisted of motion correction and phase matching between the two dual-polarity averages. The 0.4mm isotropic results are also shown after final complex-valued NLM denoising of the average. In comparison to the GRE, the 0.7mm EPI reveals less distinct signal voids as indicated in the temporal cerebellar white matter, in the posterior lobe of the cerebellum and above the sphenoidal sinus (magenta arrow). In addition, the GRE image exhibits flow artifacts (e.g. cyan arrow) and the 0.7mm EPI images appear crisper as indicated, for instance, by the more detailed depiction of white matter draining veins. Exquisite details are captured by the 0.4mm scan that are even more pronounced after denoising.

**Figure 4:**
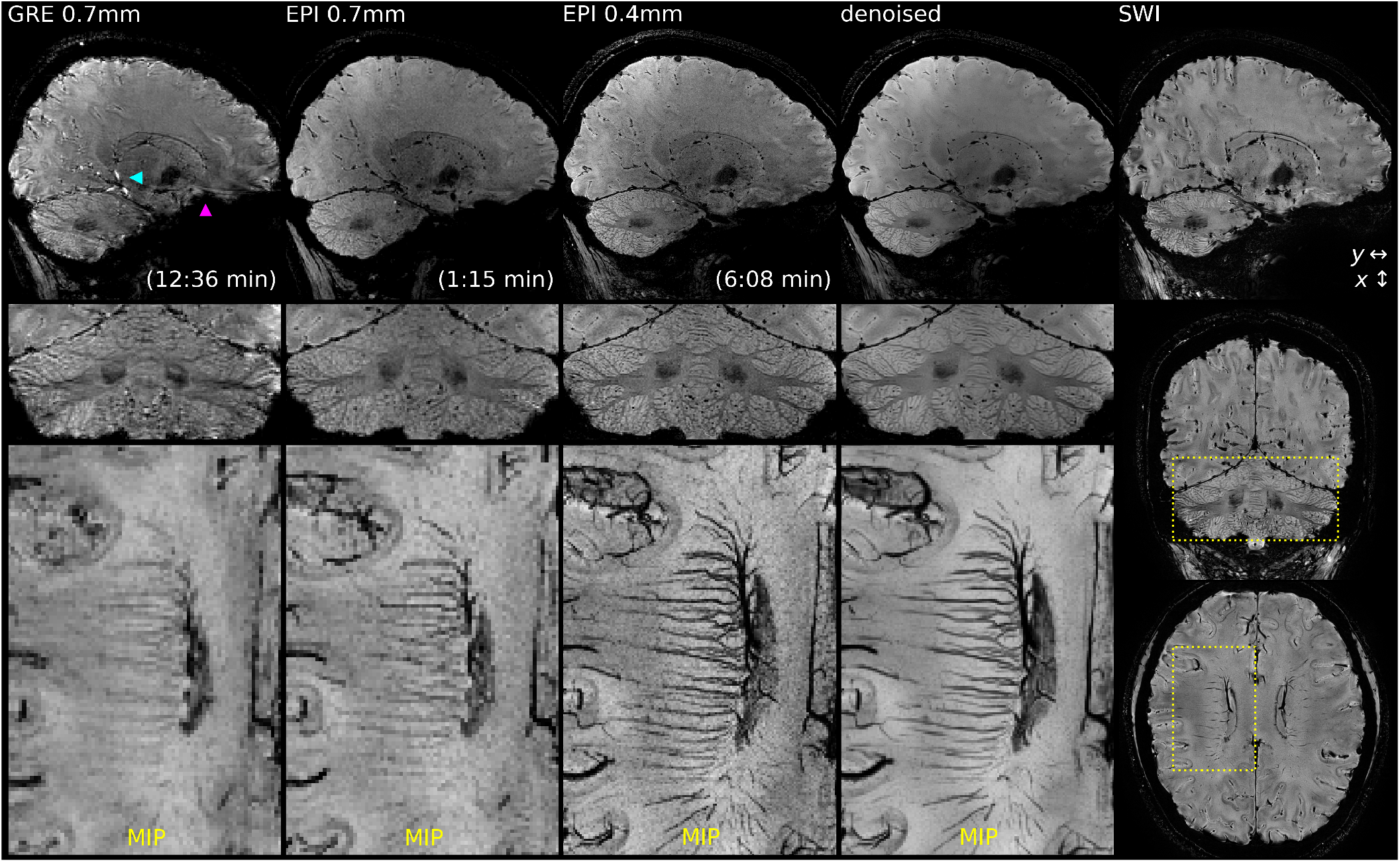
Whole-head sagittal views and magnified coronal views of the preprocessed magnitude images (top) at native resolution from single-TE experiments 1a and 2a. Right: Whole-head SWI of denoised 0.4mm EPI. Bottom: magnified axial MIPs across 5.6mm of the SWI images.

Magnified views of the corresponding quantitative susceptibility maps are shown in fig. 5. Maps at 0.4mm isotropic resolution are additionally shown with denoising prior to susceptibility mapping. Apart from more restrictive masking required for the GRE data (especially in the cerebellum), the susceptibility maps of the GRE and EPI data are nearly identical with additional fine details at 0.4mm.

**Figure 5:**
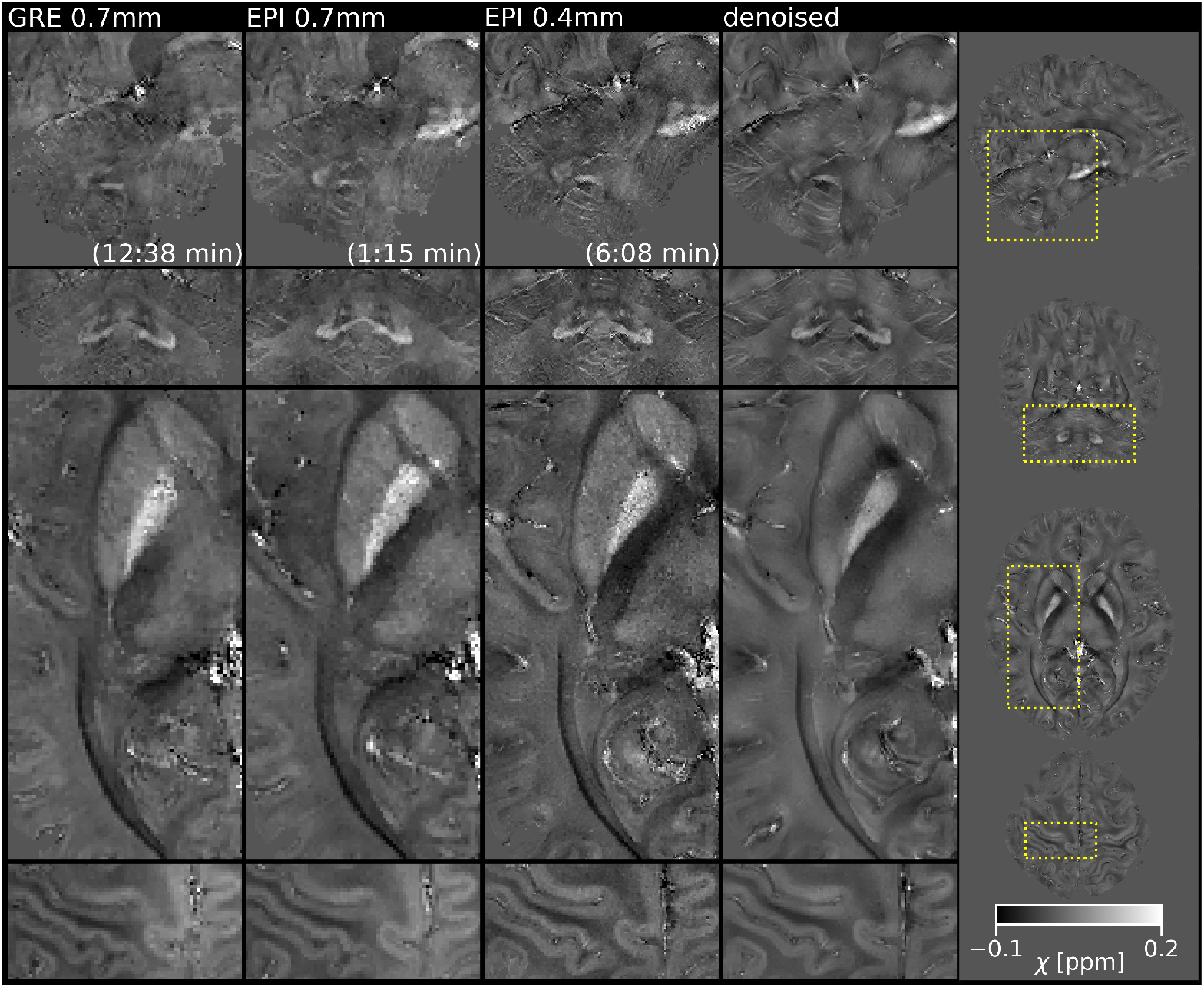
Magnified views of the susceptibility maps at native resolution from single-TE experiments 1a and 2a. The two top rows show sagittal and coronal views of the pons, red nucleus, substancia nigra, cerebellum and dentate nucleus. The two bottom rows show axial views of the striatum, the optic tract, the primary motor and somatosensory cortex. The right panel, showing whole-head views of susceptibility obtained from 0.4mm data after denoising, indicates the magnified areas.

Figure 6 shows unilateral axial views of the multi-TE magnitude images at 7T and magnified views of the corresponding SWIs and MIPs. As the 1.4mm EPI data consisted of a single measurement, only the 0.7mm EPI data underwent preprocessing, which consisted of motion correction, phase matching and distortion correction between the two ↑ & ↓ measurements per “jittered” TE. The magnified root-mean-squares combinations across TEs (RMS) are near identical for the 1.4mm EPI and GRE. In addition to the preprocessed 0.7mm RMS, the corresponding 0.7mm RMS using only ↑ phase-encoding is shown (red frame). Yellow arrows point to small structures that, accordingly, demonstrate improved actual resolution after complete preprocessing, including PF compensation.

**Figure 6:**
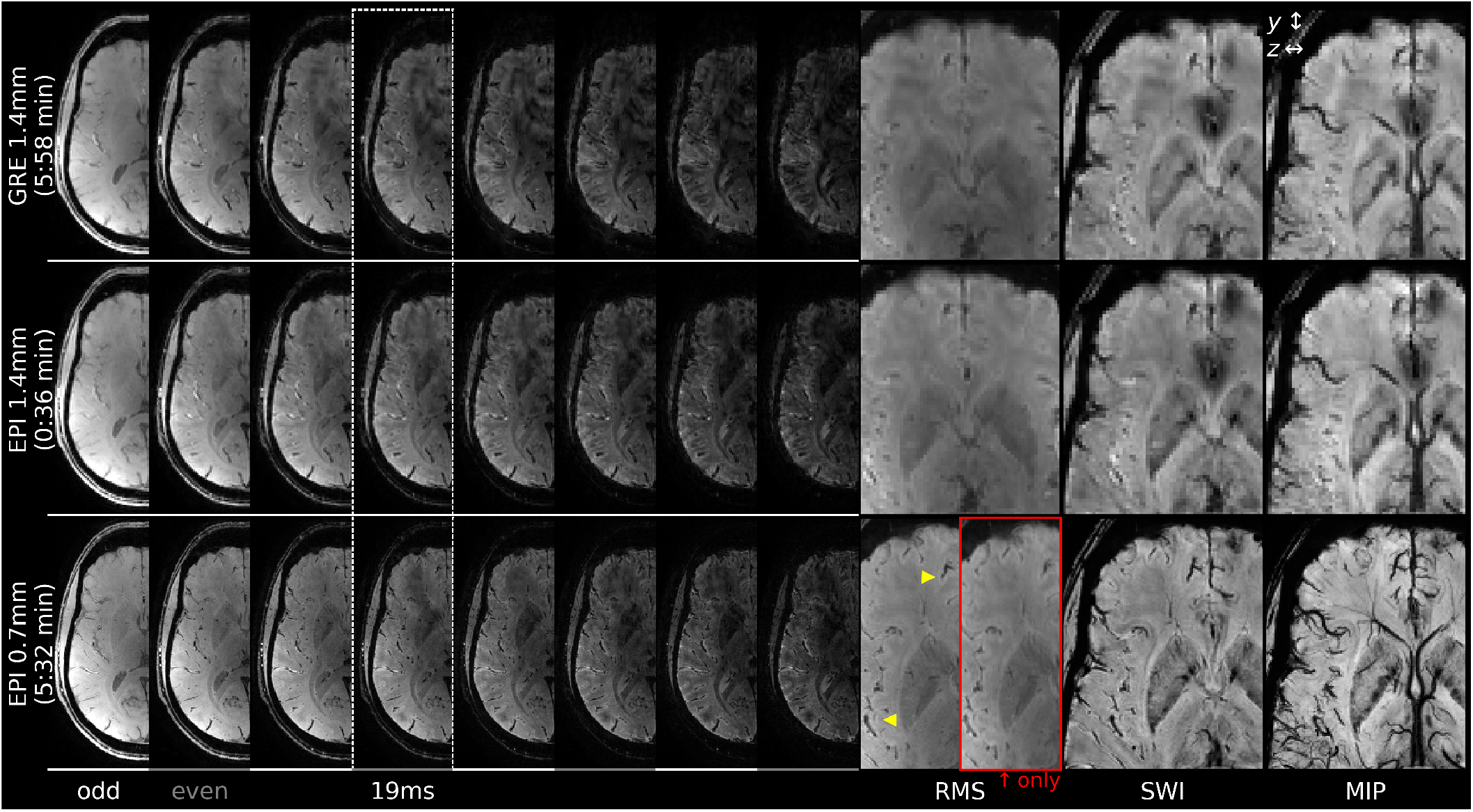
Unilateral, axial views of all TE magnitude images at native resolution from multi-TE experiments 1b (top rows, one measurement each) and 2b (bottom tow, two averages per TE, preprocessed). TE=19ms is included in all sets (dotted frame). Odd- and even-numbered measurements (interleaved sets of “jittered TEs”) are indicated by white and gray lines, respectively. Right columns: magnified axial views of the root-mean-squares combination across TEs (RMS), SWI and corresponding 5.6mm MIP. Yellow arrows indicate improved resolution after preprocessing compared to the *↑* only RMS (red frame).

Magnified views of the corresponding 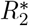 maps and quantitative susceptibility maps are shown in fig. 7. The GRE susceptibility map shows artifacts in the cerebellum (cyan arrows) and both GRE and EPI susceptibility maps at 1.4mm resolution show increased values in the axial view above the sphenoidal sinus, just like the corresponding 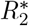 maps (magenta arrows). With EPI, eight times smaller cubic voxels (0.7mm) could be collected in about the same TA as the GRE (1.4mm). The 0.7mm data resolves more details and reduces signal dropouts, signal fluctuations and ultimately quantitative mapping errors compared to the 1.4mm GRE data.

**Figure 7:**
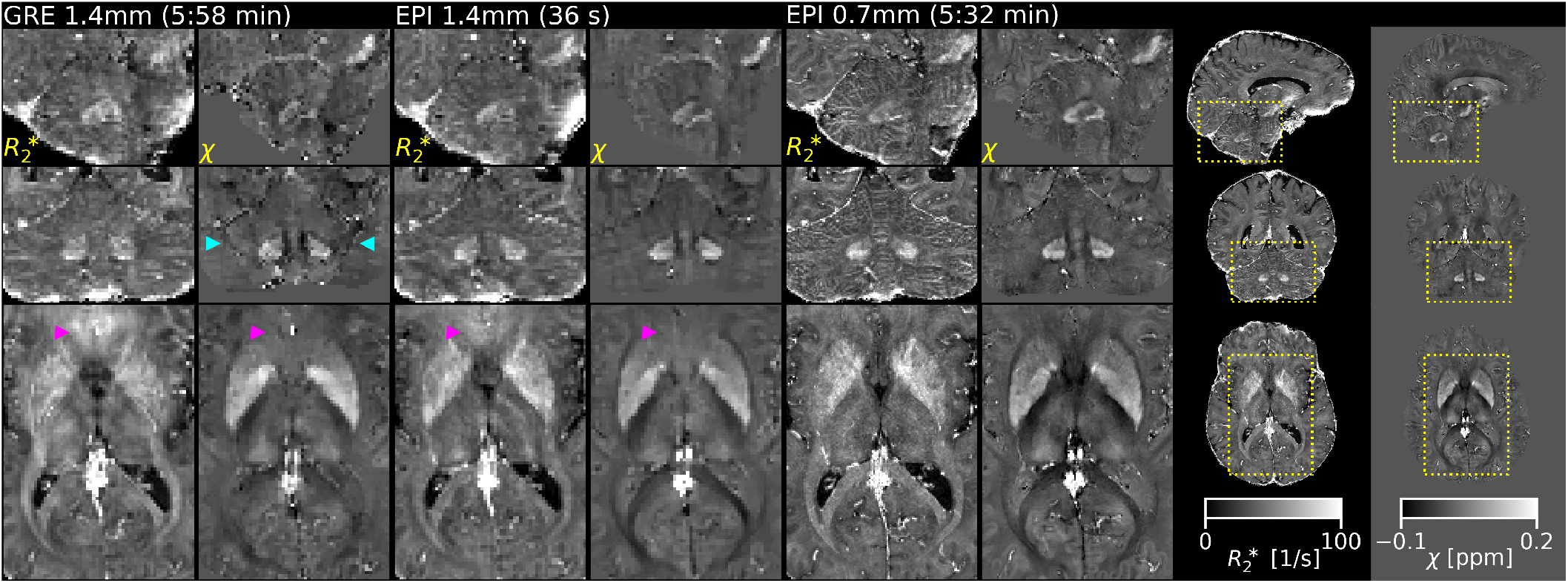
Magnified sagittal, coronal and axial views of the 7T 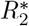 maps and susceptibility maps from multi-TE experiments 1b and 2b. Eight times smaller cubic voxels (0.7mm) in about the same TA as the GRE (1.4mm) reduce quantitative mapping errors in highly off-resonant areas (magenta arrows). The right panels, showing whole-head maps of the 0.7mm EPI, indicate the magnified areas.

Finally, fig. 8 shows unilateral axial views of the 3T multi-TE magnitude images, corresponding SWIs and MIPs, resulting 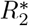 and susceptibility maps for two passes of the 0.8mm EPI protocol. Supporting Information Fig. S2 B shows that moderate motion artifacts were present in the raw images of measurements 2 and 8 and substantial motion artifacts in the raw images of measurement 7 of the second pass. Image data of measurement 7 was replaced by a copy of measurement 5 data before further preprocessing, which is equivalent to motion censoring in this context. Preprocessing of the two passes consisted of motion correction, phase matching and distortion correction before averaging, which resulted in comparable data quality demonstrated in the top two rows of fig. 8. The bottom row shows average images and maps of the same data (after motion censoring of measurement 7) without preprocessing. Detail level and overall quality of the susceptibility map without preprocessing is degraded. Notably, reduced image magnitudes, in particular for long TEs with increasing phase discrepancies between averages, result in a substantial upwards bias of 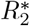 across the whole brain.

**Figure 8:**
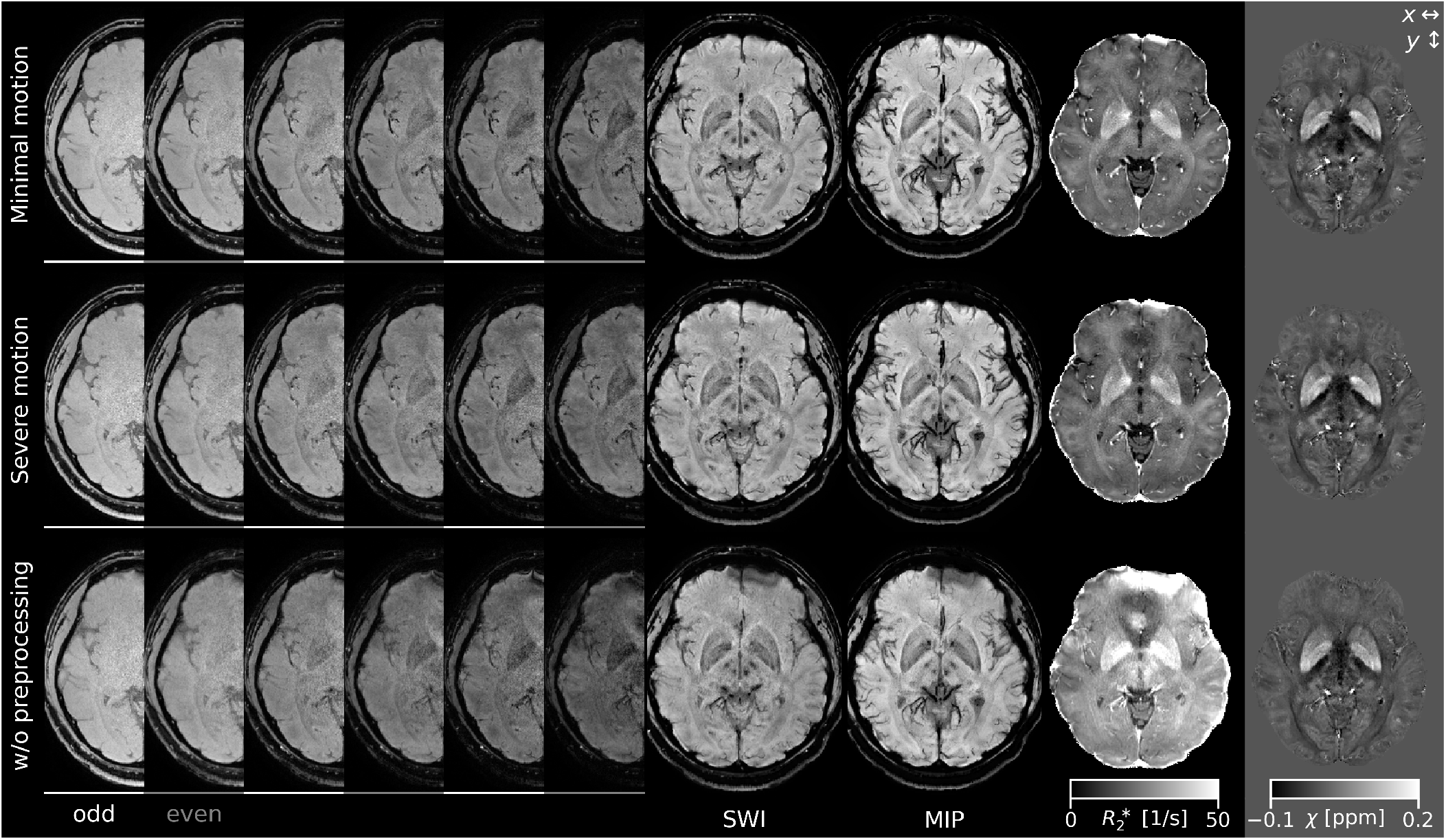
Unilateral, axial views of the magnitude images for all TEs of 3T experiment 3 (0.8mm isotropic, TA=6:22) after complete preprocessing with minimal (top row, 4 averages per TE) and with severe head motion (middle row, 3/4 averages per odd/even-numbered TE due to motion censoring of measurement 7; cf. Supporting Information Fig. S3). Bottom row: same data averaged without preprocessing (only linear phase differences in *x* direction removed). Odd- and even-numbered measurements (TEs) are indicated by white and gray lines, respectively. Right columns: axial views of the SWI and MIP, 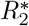 map (after denoising) and susceptibility map (without denoising).

**Figure 9:**
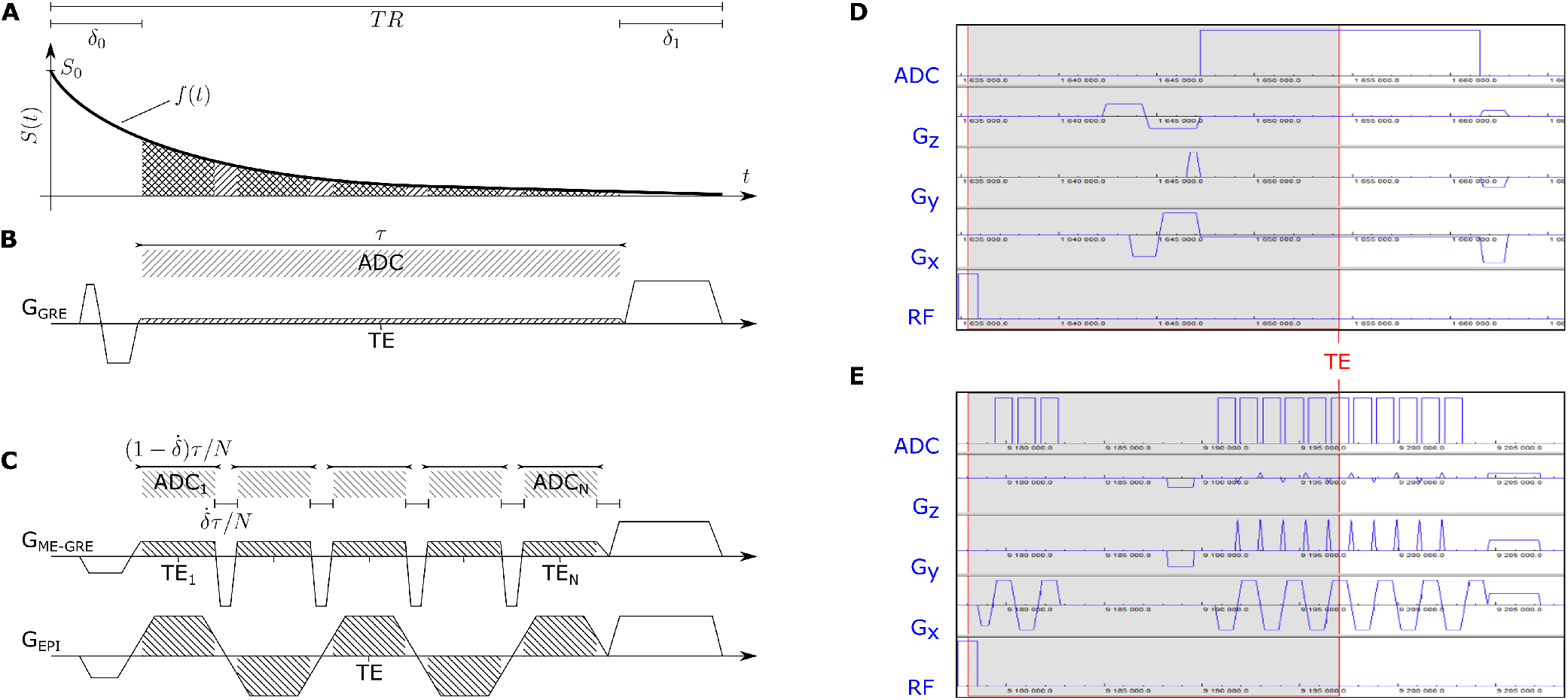
Illustration of the sampling parameters assumed for SNR efficiency calculations (A). Representative frequency-encode pulse sequences for the contiguous FID model (B; e.g. GRE with flow compensated dephaser in *δ*_0_) and for the split FID sequence model (C; monopolar multi-TE GRE or multi-shot EPI with *N* = 5 echoes each). Right: One example TR of actual pulse sequence simulations for the GRE (D) and EPI (E) of experiment 1a. ADC: analog-to-digital conversion of the sampled signal.

## 5 Discussion

### 5.1 Experimental data

The single-TE SC-EPI data of experiment 1a was effectively 36-fold accelerated (each dual-polarity measurement 72-fold) and thus about ten times faster (1:15 min) than the GRE reference (12:38 min). Magnitude images are even improved and susceptibility maps are near identical. In addition to reduced artifacts due to less intra-volume motion and field variations, no flow artifatcs are visible in the 0.7mm EPI magnitude compared to the 0.7mm GRE magnitude. For 3D-GRE imaging using partial flow compensation (only along *x* and *z* according to the UK7T protocol reference), such flow artifacts can be expected.^35^ On the other hand, segmented EPI is known to exhibit a different type of flow artifact,^36^ which was, however, not observed here. Potentially, a small amount of diffusion-weighting from the EPI frequency-encoding gradients could explain reduced blood signal.

The increased resolution of the 0.4mm SC-EPI resulted in less dropouts and showed even more remarkable details compared to the 0.7mm SC-EPI, particularly of the convoluted structures of the cerebellar cortex. The increased amount and length of the medullary veins projected by the 0.4mm susceptibility-weighted MIP also demonstrates substantially reduced partial volume effects by 1/5 of the original voxel volume. Finer details due to increased resolution and reduced sensitivity to motion with SC-EPI are also depicted in the resulting susceptibility maps of fig. 5. In particular, layers of the cortical gray matter, deep gray matter structures, cerebral white matter and even fine white matter tracts in the pons can be delineated (see Supporting Information Animation S3 for whole-head axial views).

Despite substantially increased resolution, much shorter acquisition times per measurement (*TR*_*vol*_=2:59 min) could be achieved for the ultra-highly resolved SC-EPI with 400 microns isotropic voxels compared to the 0.7mm GRE reference (TA=12:38 min). For sufficient SNR, only two averages were necessary, still resulting in less than half of the TA of the 0.7mm GRE (6:08 min). A corresponding 400 microns GRE with identical parallel imaging and ACS would be about *N/M* = 8.5 times longer (approximately 50 minutes).

A single 1.4mm isotropic multi-TE SC-EPI measurement yielded comparable data quality to the corresponding multi-TE GRE reference in about one tenth of the scan time, as demonstrated in fig. 6. In approximately the same scan time as the 1.4mm multi-TE GRE (and only half of the scan time of the 0.7mm single-TE GRE), distortion-free whole-head multi-TE SC-EPI data at 0.7mm isotropic resolution was obtained. Hereby, our implemented PF compensation restored the actual resolution, despite zero-filling in the individual measurements. Overall, significantly smaller voxels and reduced motion and field variation per measurement resulted in less signal dropouts and reduced artifacts and allows to resolve much finer details compared to the 1.4mm resolution. Furthermore, it translated to substantially improved 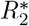 and susceptibility maps as shown in fig. 7.

Similar conclusions can be drawn from the multi-TE experiments performed at 3T, as demonstrated in fig. 8. The final averaging stage restored full phase-encode resolution due to ↑ & ↓ phase-encoding with PF compensation and, at the same time, mitigated off-resonance-induced segmentation artifacts due to dual-polarity frequency-encoding (partially in the motion-censored case). Due to particularly short acquisition times per measurement (*TR*_*vol*_=45 s), intra-volume motion sensitivity was rather small. In the presence of severe head motion (middle row), retrospective inter-volume motion correction and phase matching produced nearly similar quality as with negligible, unintentional motion (top row). Without preprocessing (bottom row), displacements and predominantly motion-induced phase discrepancies caused blurring, phase errors and severe magnitude reduction. In particular, 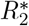 maps are substantially affected by an upwards bias, if phase matching before averaging is omitted. A single 3D-GRE acquisition of the same duration would suffer from different, presumably even stronger artifacts in the presence of similar motion. With all retrospective, image-based corrections applied, however, clear SWIs, MIPs and quantitative maps were obtained from the SC-EPI scans.

### 5.2 Retrospective corrections

The degrees of freedom through repeated measurements may be used in various ways. Here, only pragmatic, image-based processing was done, that was applicable retrospectively using readily available software.

In particular the 0.8mm multi-TE SC-EPI data, acquired at 3T over eight, already motion-insensitive measurements of only 45 s duration each, benefited from retrospective motion correction, distortion correction, phase matching, partial Fourier compensation and mitigation of (minimal) segmentation artifacts. In addition, motion censoring was feasible. Together with the other retrospective corrections, the quality of the outcome was substantially improved. This is particularly useful for study participants with difficulties to lie still (see fig. 8). The four measurements of 0.7mm multi-TE SC-EPI data acquired at 7T (1:19 min each) benefited from a similar pipeline (excluding mitigation of segmentation artfacts, see below). Even the two single-TE SC-EPI measurements at 0.7mm (33 s each) and 0.4mm isotropic resolution (2:59 min each) allowed for retrospective inter-volume motion correction and phase matching in addition to mitigating segmentation artifacts. Coregistration before averaging is particularly useful at isotropic resolutions as high as 400 microns.^37^ However, prospective intra-volume motion and phase correction would be even more beneficial. A corresponding GRE scan of about 50 minutes duration would only be feasible with prospective motion correction.

The proposed dual-polarity averaging technique has a lot in common with previously proposed Nyquist ghost correction methods, e.g. as in [^38, 39^]. One main difference is that in our approach we already start with good phase correction by applying phase navigators. Minor, residual ghosts are eliminated through dual-polarity averaging. Our main objective was the mitigation of off-resonance-induced segmentation artifacts. Nevertheless, future applications may benefit additionally from Phase Labeling for Additional Coordinate Encoding (PLACE)^39^ or Dual-Polarity GRAPPA,^40^ for instance.

We found that interleaved multi-shot EPIs with only few echo sections (e.g. 5-7 for the multi-TE experiments 1b, 2b and 3) generally do not seem to suffer as severely from off-resonance induced segmentation artifacts as the other scans, which makes dual-polarity averaging optional (cf. Supporting Information Fig. S4). This might be due to fewer off-resonance phase jumps that are further apart in k-space compared to an intermediate number of echo sections (e.g. 12 or 17 in experiments 1a and 2a). Corresponding image artifacts would be more localized and less pronounced as less phase is accumulated over shorter EPI readouts (smaller phase error tilt^16^).

In our high-resolution multi-TE scans, we used reversed phase-encoding in repeated measurements. This allowed us to compensate for PF-related resolution loss and to correct for susceptibility-induced geometric distortions in addition to inter-volume motion and phase changes. However, only small PF-related resolution loss and geometric distortions were observed in our highly-segmented images (see magnified RMS comparison to ↑ only RMS in fig. 6). The same applies to the 3T protocol. Upon distortion correction, no geometric mismatch with a conventional *T*_1_-weighted anatomical scan was visible (see Supporting Information Fig. S5). At 3T, both dual-polarity frequency-encoding and ↑ & ↓ phase-encoding were applied, resulting in eight measurements in total (four per TE set).

The reversed phase-encoding approach is closely related to the 3D blip-up/blip-down acquisition (BUDA) that has recently been utilized successfully for QSM^41^ and 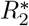 mapping.^42^ BUDA can potentially take even more advantage of the scanning speed by acquiring complementary sampling patterns per blip-up and blip-down shot. Compared to BUDA applications proposed thus far, this work relies on moderate parallel imaging undersampling for both phase-encode directions and rather small distortions at much higher spatial resolutions, as facilitated through strong interleaved multishot segmentation. The use of skipped-CAIPI sampling in each self-contained blip-up and blip-down image allowed us to perform corrections retrospectively on fully reconstructed images instead of a joint reconstruction, where image-based corrections may become more difficult. Still, future applications could also benefit from advanced BUDA acquisition and image reconstruction.

Importantly, the image-based, retrospective correction strategy followed here makes the proposed acquisition and correction approach very accessible. The use of CAIPIRINHA sampling is not even mandatory with reduced undersampling,^10, 11^ and therefore, it is even compatible with more basic image reconstruction, provided some sort of interleaved multi-shot 3D-EPI sequence with adequate magnitude and phase reconstruction is available. Even without retrospective corrections and averaging, 1mm isotropic or anisotropic SWI or 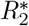 mapping well below one minute scan time could be performed in clinical settings. In research settings, more elaborate methods or improved hardware, including parallel transmission flip angle homogenization^43^ or more powerful imaging gradient systems,^44^ can be used in addition or alternatively without diminishing our main findings.

## 6 Conclusions

Interleaved multi-shot 3D-EPI is highly SNR-efficient and well suited for high-resolution single-TE and multi-TE gradient echo imaging at high and ultra-high fields. Single scans are acquired rapidly, making them relatively motion-insensitive and providing many degrees of freedom to further increase robustness against experimental imperfections over repeated short measurements. Therefore, interleaved multi-shot 3D-EPI, with or without CAIPIRINHA sampling, readily allows for rapid and robust QSM and 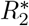 mapping at 7T and 3T.

## Supporting information

Supporting Information Animation S3

## Data availability statement

Separate nipype pipelines for basic and for extended preprocessing are shared under https://github.com/mrphysics-bonn/ep3d-highres-preprocessing (short hash at time of submission: xxxxxxxx). The skipped-CAIPI 3D-EPI sequence can be accessed via the Siemens C2P exchange platform at https://webclient.eu.api.teamplay.siemens-healthineers.com/c2p (v2.0 at time of submission).

## Acknowledgments

We want to thank Mohammad Shahid for the nipype implemention of the extended preprocessing pipeline for the Rhineland study.

## Supporting information

The following supporting information is available as part of the online article:

**Figure S1**. Numerical phantom simulations of dualpolarity frequency-encoding for increasing off-resonance frequencies, Δ*f* (96 phase-encode lines, segmentation factor *S* = 8, EPI factor = echo sections *N* = 96*/S* = 12). Segmentation artifacts are clearly visible in the individual dual-polarity images (+ and -) for 440Hz and 880Hz off-resonance frequencies that are mitigated through dual-polarity averaging. Additional, massive off-resonance aliasing without echo time shifting (w/o ETS) does not disappear through dual-polarity averaging (not shown).

**Figure S2**. Axial SC-EPI magnitude views with “jit-tererd” multi-TE measurements of two passes of the 3T Rhineland Study 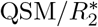 protocol. The EPI trajectories displayed to the left of each rephased multi-TE set indicate the respective frequency (*k*_*x*_) and phase-encode (*k*_*y*_) polarity. The first protocol pass (A) has been acquired without substantial head motion. During the second protocol pass (B), the subject was instructed to reposition the legs once (implying realistic motion) and to nod with the head once strongly (exaggerated motion). Measurements 2 (orange frame) and 7 (red frame) were affected, respectively. Apart from involuntary motion artifacts in measurement 8 (orange frame), all other scans were found normal. Measurement 7 was replaced by a copy of measurement 5 before preprocessing. This corresponds to motion censoring, as the copy of measurement 5 does not add information. Consequently, for the three respective TEs, only partial mitigation of segmentation artifacts is achieved (complete in ↑ phase-encoding, none in ↓ phase-encoding). Still, results after preprocessing are comparable to the minimal motion pass, although slightly reduced in SNR (see Fig. 8 in main text).

**Animation S3**. Axial views across the whole head at 400 microns isotropic. SWIs, MIPs (across 5.6mm) and susceptibility maps are based on NLM-denoised SC-EPI data.

**Figure S4**. Raw images vs. average images demonstrate mitigation of off-resonance-induced segmentation artifacts in all dual-polarity SC-EPI scans at 7T using medium large EPI factors (rows 1-2), and negligible segmentation artifacts in multi-TE scans at 7T (rows 3-4) and 3T (rows 5-6) using small EPI factors. Slices that should show most prominent off-resonance artifacts were selected. Yet, clear segmentation artifacts are only visible in rows 1-2 (stripe patterns, framed yellow and magnified). The fourth column additionally shows the shortest TE of all multi-TE images as a guidance to distinguish reduced signal-dropouts from other off-resonance artifacts.

**Figure S5**. SC-EPI average magnitude images at the shortest TE compared to a conventional T1-weighted anatomical image (T1w, native isotropic resolution: 1mm in rows 1-2, 0.8mm in rows 3-5). Gray/white matter boundaries were derived from the T1w by thresholding after coregistration (mutual information) and interpolation (sinc) to the SC-EPI. The SC-EPIs of rows 1-3 did not undergo geometric distortion correction. Yet, minor geometric mismatch to T1w is only apparent in the magnified views of row 2 and 3 (third column).

## Appendices

### A SNR-efficient sampling of a free-induction decay

Regardless of the GRE sequence type, sampling of a single free-induction decay (FID) can be considered. The SNR is proportional to the Fourier series DC coefficient divided by the noise standard deviation.^45^

Figure 9A,B defines the sequence timing assumed for the FID model. Over the sampling window, *τ* (*TR*), the steady-state signal *S*_0_ = *S*(*t* = 0, *TR, α*) shall be weighted by a generic signal envelope, *f* (*t*). Assuming Gaussian noise with a variance of *σ*^2^ yields the following SNR equation:

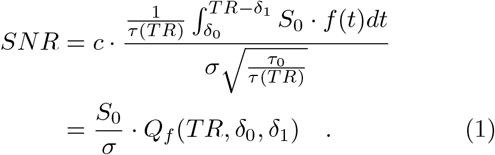

The receive bandwidth factor 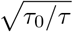 scales the noise standard deviation (*τ*_0_: arbitrary normalization) and the global factor *c* accounts for all other factors that scale the signal magnitude. These factors are incorporated in the “quality factor”^45^ together with the cumulative signal over the sampling window:

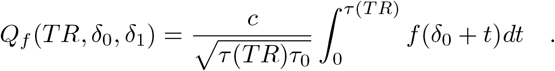

The quality factor of a monoexponential 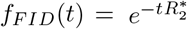 is

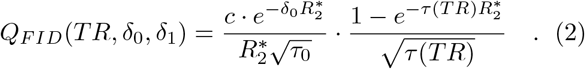

Substituting the steady-state signal using the Ernst angle,

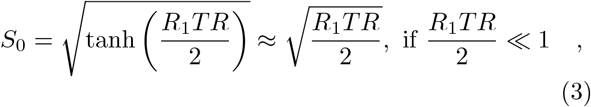

into eq. (1), the sampled FID SNR efficiency becomes:

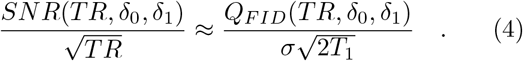

Solving 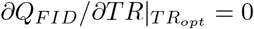 yields

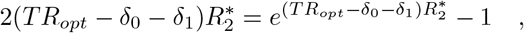

which is satisfied by the efficiency-optimal *TR*_*opt*_

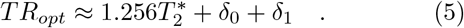

The peak efficiency is obtained by substituting *Q*_*F ID*_ according to eq. (2) with *TR*_*opt*_ into eq. (4):

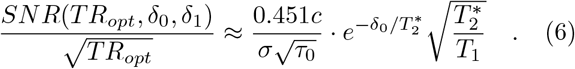

### B SNR-efficient sampling of ME-GRE-type sequences

If, according to fig. 9 C, we assume a fraction 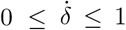 of the FID sampling window interspersed over a split sampling window of reduced cumulative time, 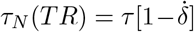, instead of eq. (2) one finds

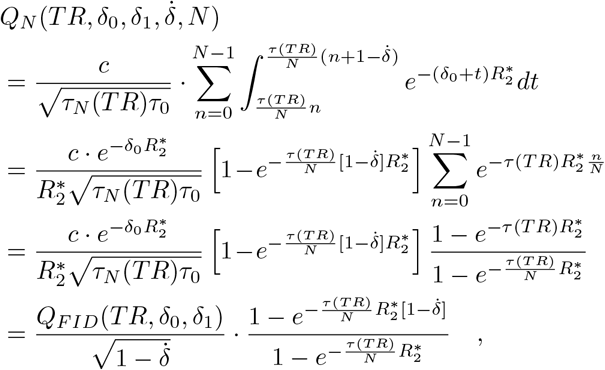

which has the coherent special cases:

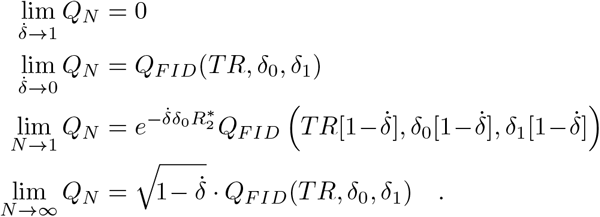

Corresponding limits apply to the SNR efficiency:

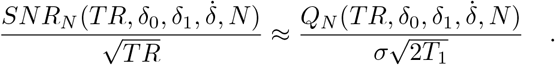

The first two cases are trivial. The third case corresponds to a single signal acquisition interval, just like the FID model. However, the acquisition window is shorter by 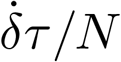 (fig. 9 B,C). The efficiency-optimal 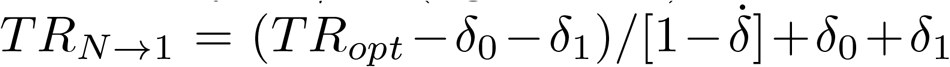 is increased accordingly, but the peak SNR efficiency is identical to the FID model.

The last case is the upper limit for many acquisition intervals (e.g. single-shot EPI). The efficiency-optimal 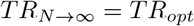 is the same as for the FID. However, the peak SNR efficiency is scaled down by 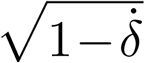.

Figure 9 D and E depict actual pulse sequence examples for the FID model and the multi-echo model, respectively. The examples correspond to the 0.7mm single-TE GRE and EPI protocols of experiment 1a.

## References

[1] Langkammer Christian, Schweser Ferdinand, Krebs Nikolaus, et al. Quantitative Susceptibility Mapping (QSM) as a Means to Measure Brain Iron? A Post Mortem Validation Study. NeuroImage. 2012;62(3):1593–1599.

[2] Hametner Simon, Endmayr Verena, Deistung Andreas, et al. The Influence of Brain Iron and Myelin on Magnetic Susceptibility and Effective Transverse Relaxation - A Biochemical and Histological Validation Study. NeuroImage. 2018;179:117–133.

[3] Stüber Carsten, Morawski Markus, Schäfer Andreas, et al. Myelin and Iron Concentration in the Human Brain: A Quantitative Study of MRI Contrast. NeuroImage. 2014;93:95–106.

[4] Chen Jingjia, Gong Nan-Jie, Chaim Khallil Taverna, Otaduy Maria Concepción García, Liu Chunlei. Decompose Quantitative Susceptibility Mapping (QSM) to Sub-Voxel Diamagnetic and Paramagnetic Components Based on Gradient-Echo MRI Data. NeuroImage. 2021;242:118477.

[5] Dimov Alexey V., Gillen Kelly M., Nguyen Thanh D., et al. Magnetic Susceptibility Source Separation Solely from Gradient Echo Data: Histological Validation. Tomography. 2022;8(3):1544–1551.

[6] Dimov Alexey V., Nguyen Thanh D., Gillen Kelly M., et al. Susceptibility Source Separation from Gradient Echo Data Using Magnitude Decay Modeling. Journal of Neuroimaging. 2022;32(5):852–859.

[7] Bilgic Berkin, Xie Luke, Dibb Russell, et al. Rapid Multi-Orientation Quantitative Susceptibility Mapping. NeuroImage. 2016;125:1131–1141.

[8] Sun Hongfu, Wilman Alan H.. Quantitative Susceptibility Mapping Using Single-Shot Echo-Planar Imaging. Magnetic Resonance in Medicine. 2015;73(5):1932–1938.

[9] Langkammer Christian, Bredies Kristian, Poser Benedikt A., et al. Fast Quantitative Susceptibility Mapping Using 3D EPI and Total Generalized Variation. NeuroImage. 2015;111:622–630.

[10] Sati P., Thomasson D. M., Li N., et al. Rapid, High-Resolution, Whole-Brain, Susceptibility-Based MRI of Multiple Sclerosis. Multiple Sclerosis Journal. 2014;20(11):1464–1470.

[11] Zwanenburg Jaco J.M., Versluis Maarten J., Luijten Peter R., Petridou Natalia. Fast High Resolution Whole Brain T2* Weighted Imaging Using Echo Planar Imaging at 7T. NeuroImage. 2011;56(4):1902–1907.

[12] Wang Difei, Ehses Philipp, Stöcker Tony, Stirnberg Rüdiger. Reproducibility of Rapid Multiparameter Mapping at 3T and 7T with Highly Segmented and Accelerated 3D-EPI. Magnetic Resonance in Medicine. 2022;(June):1–16.

[13] Marques José P., Khabipova Diana, Gruetter Rolf. Studying Cyto and Myeloarchitecture of the Human Cortex at Ultra-High Field with Quantitative Imaging: R1, R2* and Magnetic Susceptibility. NeuroImage. 2017;147:152–163.

[14] Breuer Felix A., Blaimer Martin, Mueller Matthias F., et al. Controlled Aliasing in Volumetric Parallel Imaging (2D CAIPIRINHA). Magnetic Resonance in Medicine. 2006;55(3):549–556.

[15] Stirnberg Rüdiger, Stöcker Tony. Segmented K-space Blipped-controlled Aliasing in Parallel Imaging for High Spatiotemporal Resolution EPI. Magnetic Resonance in Medicine. 2021;85(3):1540–1551.

[16] Feinberg DA, Oshio Koichi. Phase Errors in Multi-Shot Echo Planar Imaging. Magnetic resonance in medicine. 1994;32(4):535–539.

[17] Jellú?s Vladimír, Kannengiesser Stephan A R. Adaptive Coil Combination Using a Body Coil Scan as Phase Reference. In: Proceedings of the International Society of Magnetic Resonance in Medicine, vol. 23: ; 2014.

[18] Rua Catarina, Clarke William T., Driver Ian D., et al. Multi-Centre, Multi-Vendor Reproducibility of 7T QSM and R2* in the Human Brain: Results from the UK7T Study. NeuroImage. 2020;223(April):117358.

[19] Stirnberg Rüdiger, Brenner Daniel, Stöcker Tony, Shah N. Jon. Rapid Fat Suppression for Three-Dimensional Echo Planar Imaging with Minimized Specific Absorption Rate. Magnetic Resonance in Medicine. 2016;76(5):1517–1523.

[20] Clarke W. UK7T Network Harmonized Neuroimaging Protocols https://ora.ox.ac.uk/objects/uuid:55ca977f-62df-4cbf-b300-2dc893e36647 x(accessed 2023-09-13); 2018.

[21] Ivanov Dimu, Barth Markus, Uludağ. Kâmil, Poser Benedikt A.. Robust ACS Acquisition for 3D Echo Planar Imaging. In: Proceedings of the International Society of Magnetic Resonance in Medicine, vol. 23: :2059–2059; 2015.

[22] Breteler Monique M.B., Stöcker Tony, Pracht Eberhard, Brenner Daniel, Stirnberg Rüdiger. MRI in the Rhineland Study: A Novel Protocol for Population Neuroimaging. Alzheimer’s & Dementia. 2014;10(4):P92–P92.

[23] Jenkinson Mark, Beckmann Christian F., Behrens Timothy E J, Woolrich Mark W., Smith Stephen M.. Fsl. NeuroImage. 2012;62(2):782–790.

[24] Koopmans Peter J., Pfaffenrot Viktor. Enhanced POCS Reconstruction for Partial Fourier Imaging in Multi-Echo and Time-Series Acquisitions. Magnetic Resonance in Medicine. 2021;85(1):140–151.

[25] Manjón José V., Coupé Pierrick, Martí-Bonmatí Luis, Collins D. Louis, Robles Montserrat. Adaptive Non-Local Means Denoising of MR Images with Spatially Varying Noise Levels. Journal of Magnetic Resonance Imaging. 2010;31(1):192–203.

[26] Tustison Nicholas J., Cook Philip A., Holbrook Andrew J., et al. The ANTsX Ecosystem for Quantitative Biological and Medical Imaging. Scientific Reports. 2021;11(1):9068.

[27] Haacke E. Mark, Xu Yingbiao, Cheng Yu-Chung N., Reichenbach Jürgen R.. Susceptibility Weighted Imaging (SWI). Magnetic Resonance in Medicine. 2004;52(3):612–618.

[28] Eckstein Korbinian, Bachrata Beata, Hangel Gilbert, et al. Improved Susceptibility Weighted Imaging at Ultra-High Field Using Bipolar Multi-Echo Acquisition and Optimized Image Processing: CLEAR-SWI. NeuroImage. 2021;237:118175.

[29] Abdul-Rahman Hussein S, Gdeisat Munther, Burton David R, Lalor Michael J, Lilley Francis, Moore Christopher J. Fast and Robust Three-Dimensional Best Path Phase Unwrapping Algorithm.. Applied optics. 2007;46(26):6623–6635.

[30] Schweser Ferdinand, Deistung Andreas, Lehr Berengar Wendel, Reichenbach Jürgen Rainer. Quantitative Imaging of Intrinsic Magnetic Tissue Properties Using MRI Signal Phase: An Approach to in Vivo Brain Iron Metabolism?. NeuroImage. 2011;54(4):2789–2807.

[31] Wu Bing, Li Wei, Guidon Arnaud, Liu Chunlei. Whole Brain Susceptibility Mapping Using Compressed Sensing. Magnetic Resonance in Medicine. 2012;67(1):137–147.

[32] Schweser Ferdinand, Sommer Karsten, Deistung Andreas, Reichenbach Jürgen Rainer. Quantitative Susceptibility Mapping for Investigating Subtle Susceptibility Variations in the Human Brain.. NeuroImage. 2012;62(3):2083–100.

[33] Deistung Andreas, Schweser Ferdinand, Reichenbach Jürgen R.. Overview of Quantitative Susceptibility Mapping. NMR in Biomedicine. 2017;30(4):e3569.

[34] Hagberg G.e., Indovina I., Sanes J.n., Posse S.. Real-Time Quantification of T2* Changes Using Multiecho Planar Imaging and Numerical Methods. Magnetic Resonance in Medicine. 2002;48(5):877–882.

[35] Deistung Andreas, Dittrich Enrico, Sedlacik Jan, Rauscher Alexander, Reichenbach Jürgen R.. ToF-SWI: Simultaneous Time of Flight and Fully Flow Compensated Susceptibility Weighted Imaging. Journal of Magnetic Resonance Imaging. 2009;29(6):1478–1484.

[36] Beck Gabriele, Li Debiao, Mark Haacke E., Noll Tobias G., Schad Lothar R.. Reducing Oblique Flow Effects in Interleaved EPI with a Centric Reordering Technique. Magnetic Resonance in Medicine. 2001;45(4):623–629.

[37] Lüsebrink Falk, Sciarra Alessandro, Mattern Hendrik, Yakupov Renat, Speck Oliver. T1-Weighted in Vivo Human Whole Brain MRI Dataset with an Ultrahigh Isotropic Resolution of 250 M m. Scientific Data. 2017;4(1):170032.

[38] Chen Nan-kuei, Wyrwicz Alice M.. Removal of EPI Nyquist Ghost Artifacts with Two-Dimensional Phase Correction. Magnetic Resonance in Medicine. 2004;51(6):1247–1253.

[39] Xiang Qing-San, Ye Frank Q.. Correction for Geometric Distortion and N/2 Ghosting in EPI by Phase Labeling for Additional Coordinate Encoding (PLACE). Magnetic Resonance in Medicine. 2007;57(4):731–741.

[40] Hoge W. Scott, Polimeni Jonathan R.. Dual-Polarity GRAPPA for Simultaneous Reconstruction and Ghost Correction of Echo Planar Imaging Data. Magnetic Resonance in Medicine. 2016;76(1):32–44.

[41] Bilgic Berkin, Poser Benedikt A., Langkammer Christian, Setsompop Kawin, Liao Congyu. 3D-BUDA Enables Rapid Distortion-Free QSM Acquisition. In: Proceedings of the International Society of Magnetic Resonance in Medicine:0596; 2020.

[42] Chen Zhifeng, Liao Congyu, Cao Xiaozhi, et al. 3D-EPI Blip-up/down Acquisition (BUDA) with CAIPI and Joint Hankel Structured Low-Rank Reconstruction for Rapid Distortion-Free High-Resolution T2* Mapping. Magnetic Resonance in Medicine. 2023;89(5):1961–1974.

[43] Löwen Daniel, Pracht Eberhard D, Stirnberg Rüdiger, Liebig Patrick, Stöcker Tony. Interleaved Binomial kT-Points for Water-selective Imaging at 7T. Magnetic Resonance in Medicine. 2022;:1–10.

[44] Feinberg David A., Beckett Alexander J. S., Vu An T., et al. Next-Generation MRI Scanner Designed for Ultra-High-Resolution Human Brain Imaging at 7 Tesla. Nature Methods. 2023;:1–10.

[45] Deichmann Ralf, Adolf Holger, Nöth Ulrike, Kuchenbrod Erwin, Schwarzbauer Christian, Haase Axel. Calculation of Signal Intensities in Hybrid Sequences for Fast NMR Imaging. Magnetic Resonance in Medicine. 1995;34(3):481–489.

