## Supporting Information for "Rapid submillimeter QSM and R_2_^*^ mapping using interleaved multi-shot 3D-EPI at 7 and 3 Tesla"

### Supporting Information Figures for: Rapid submillimeter QSM and $R_2^*$ mapping using interleaved multi-shot 3D-EPI at 7 and 3 Tesla

Rüdiger Stirnberg[1], Andreas Deistung[2], Jürgen R. Reichenbach[3], Monique M. B. Breteler[4,5], and Tony Stöcker[1,6]

[1] *MR Physics, German Center for Neurodegenerative Diseases (DZNE), Bonn, Germany*
[2] *Clinic and Outpatient Clinic for Radiology, University Hospital Halle (Saale), University Medicine Halle, Halle (Saale), Germany*
[3] *Medical Physics Group, Institute of Diagnostic and Interventional Radiology, Jena University Hospital, Jena, Germany*
[4] *Population Health Sciences, German Center for Neurodegenerative Diseases (DZNE), Bonn, Germany*
[5] *Faculty of Medicine, Institute for Medical Biometry, Informatics and Epidemiology (IMBIE), University of Bonn, Bonn, Germany*
[6] *Department of Physics & Astronomy, University of Bonn, Bonn, Germany*

December 29, 2023

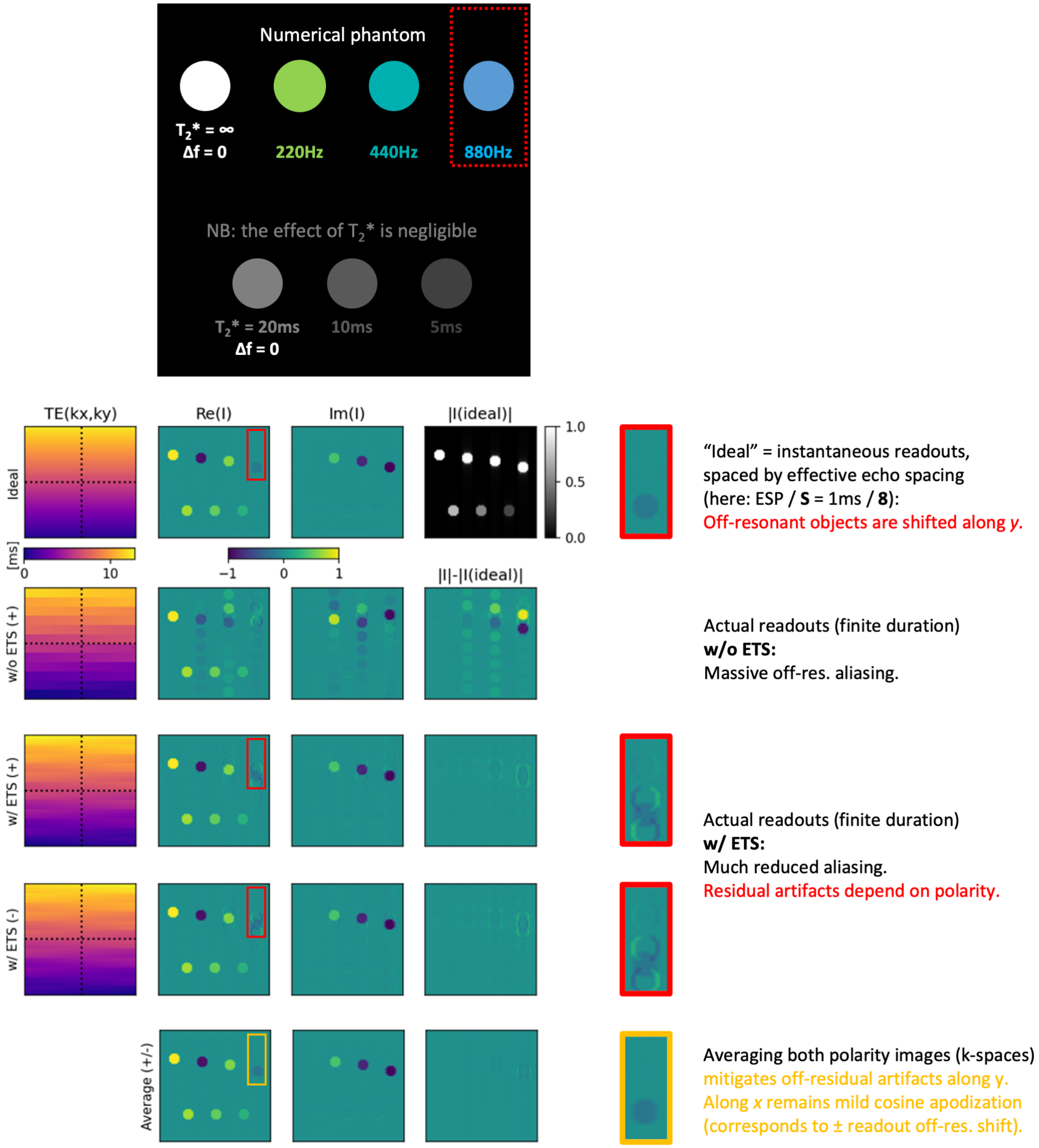


**Figure S1.** Numerical phantom simulations of dual-polarity frequency-encoding for increasing off-resonance frequencies, $\Delta f$ (96 phase-encode lines, segmentation factor $S = 8$, EPI factor = echo sections $N = 96/S = 12$). Segmentation artifacts are clearly visible in the individual dual-polarity images (+ and -) for 440Hz and 880Hz off-resonance frequencies that are mitigated through dual-polarity averaging. Additional, massive off-resonance aliasing without echo time shifting (w/o ETS) does not disappear through dual-polarity averaging (not shown).

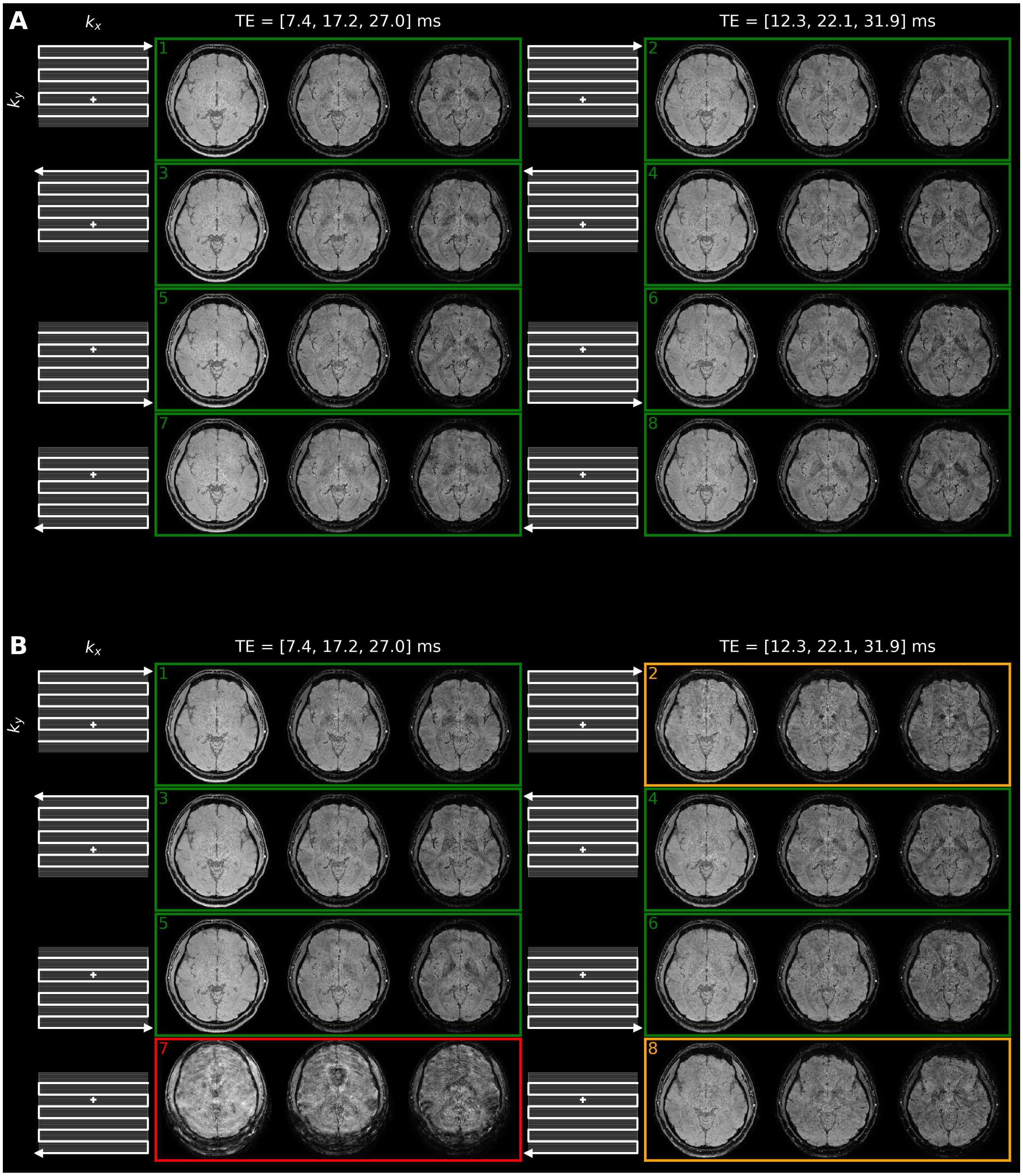


**Figure S2.** Axial SC-EPI magnitude views with "jittererd" multi-TE measurements of two passes of the 3T Rhineland Study QSM/$R_2^*$ protocol. The EPI trajectories displayed to the left of each rephased multi-TE set indicate the respective frequency ($k_x$) and phase-encode ($k_y$) polarity. The first protocol pass (A) has been acquired without substantial head motion. During the second protocol pass (B), the subject was instructed to reposition the legs once (implying realistic motion) and to nod with the head once strongly (exaggerated motion). Measurements 2 (orange frame) and 7 (red frame) were affected, respectively. Apart from involuntary motion artifacts in measurement 8 (orange frame), all other scans were found normal. Measurement 7 was replaced by a copy of measurement 5 before preprocessing. This corresponds to motion censoring, as the copy of measurement 5 does not add information. Consequently, for the three respective TEs, only partial mitigation of segmentation artifacts is achieved (complete in ↑ phase-encoding, none in ↓ phase-encoding). Still, results after preprocessing are comparable to the minimal motion pass, although slightly reduced in SNR (see Fig. 8 in main text).

**Animation S3.** Axial views across the whole head at 400 microns isotropic. SWIs, MIPs (across 5.6mm) and susceptibility maps are based on NLM-denoised SC-EPI data.

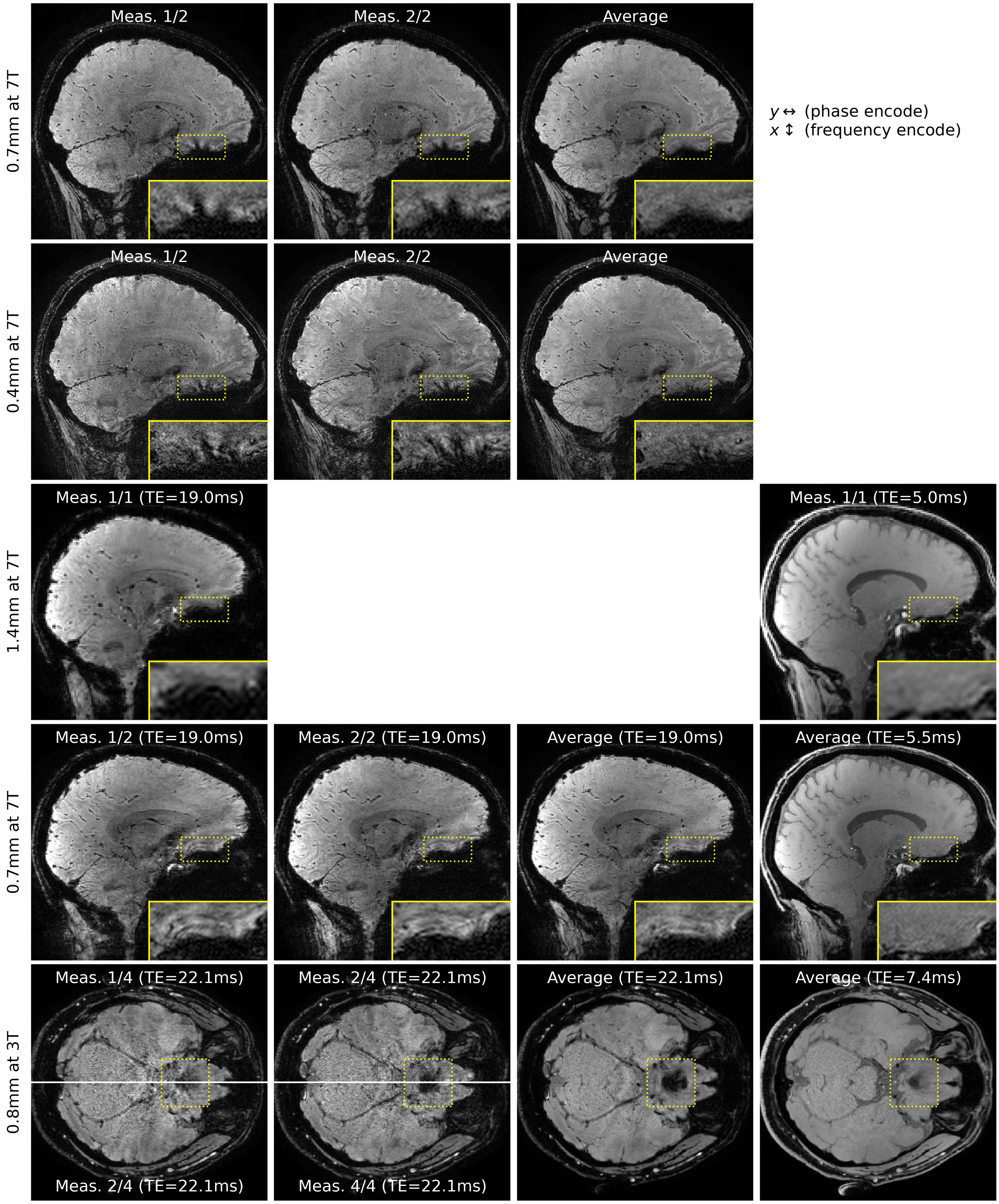


**Figure S4.** Raw images vs. average images demonstrate mitigation of off-resonance-induced segmentation artifacts in all dual-polarity SC-EPI scans at 7T using medium large EPI factors (rows 1-2), and negligible segmentation artifacts in multi-TE scans at 7T (rows 3-4) and 3T (rows 5-6) using small EPI factors. Slices that should show most prominent off-resonance artifacts were selected. Yet, clear segmentation artifacts are only visible in rows 1-2 (stripe patterns, framed yellow and magnified). The fourth column additionally shows the shortest TE of all multi-TE images as a guidance to distinguish reduced signal-dropouts from other off-resonance artifacts.

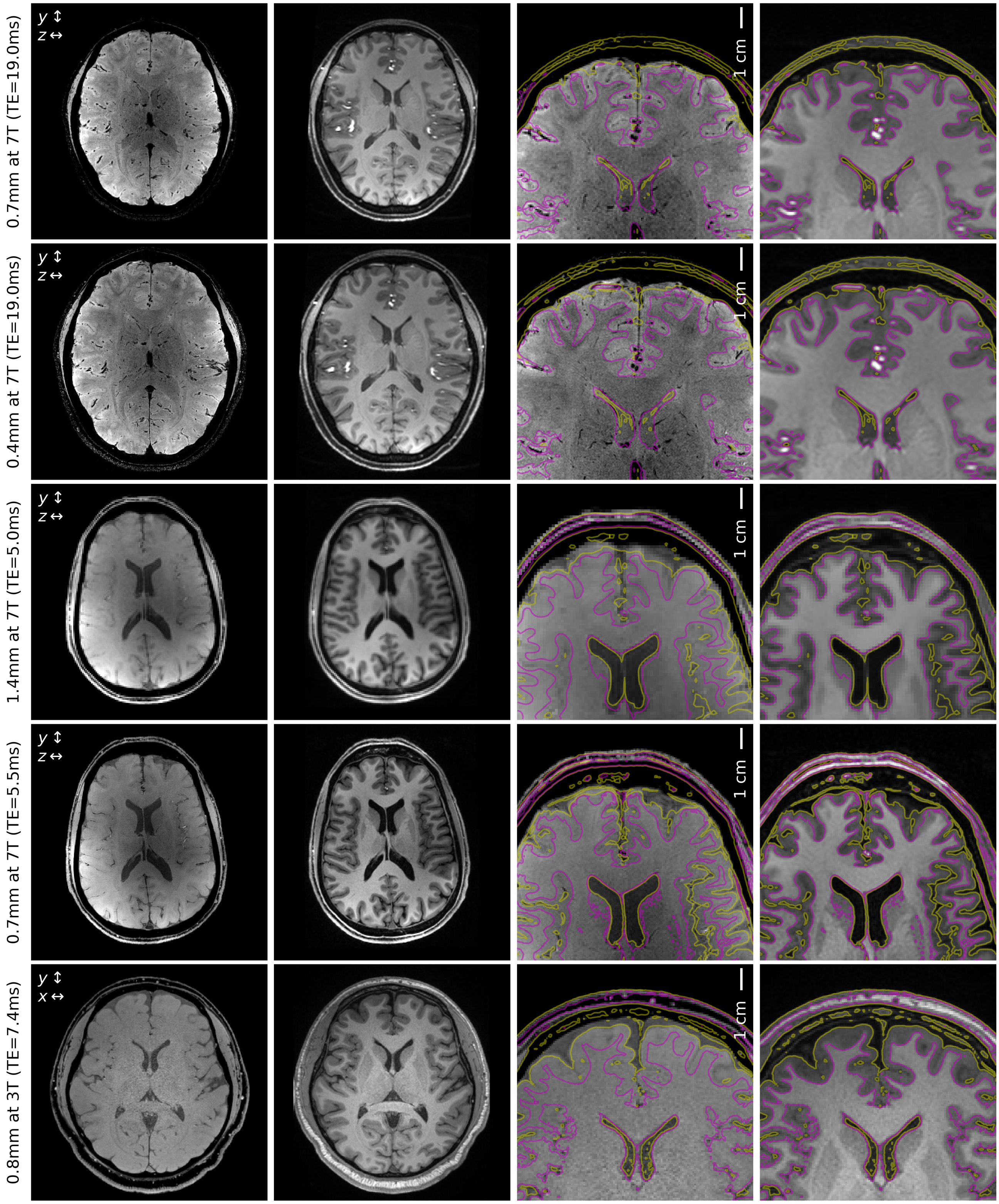


**Figure S5.** SC-EPI average magnitude images at the shortest TE compared to a conventional T1-weighted anatomical image (T1w, native isotropic resolution: 1mm in rows 1-2, 0.8mm in rows 3-5). Gray/white matter boundaries were derived from the T1w by thresholding after coregistration (mutual information) and interpolation (sinc) to the SC-EPI. The SC-EPIs of rows 1-3 did not undergo geometric distortion correction. Yet, minor geometric mismatch to T1w is only apparent in the magnified views of row 2 and 3 (third column).
